# Co-development of gut microbial metabolism and visual neural circuitry over human infancy

**DOI:** 10.1101/2024.07.24.24310884

**Authors:** Kevin S. Bonham, Emma T. Margolis, Guilherme Fahur Bottino, Ana Sobrino, Fadheela Patel, Shelley McCann, Michal R. Zieff, Marlie Miles, Donna Herr, Lauren Davel, Cara Bosco, Khula South Africa Data Collection Team, Curtis Huttenhower, Nicolò Pini, Daniel C. Alexander, Derek K. Jones, Steve C. R. Williams, Dima Amso, Melissa Gladstone, William P. Fifer, Kirsten A. Donald, Laurel J. Gabard-Durnam, Vanja Klepac-Ceraj

## Abstract

Infancy is a time of elevated neuroplasticity supporting rapid brain and sensory development. The gut microbiome, also undergoing extensive developmental changes in early life, may influence brain development through metabolism of neuroactive compounds. Here, we leverage longitudinal data from 194 infants across the first 18 months of life to show that microbial genes encoding enzymes that metabolize molecules playing a key role in modulating early neuroplasticity are associated with visual cortical neurodevelopment, measured by the Visual-Evoked Potential (VEP). Neuroactive compounds included neurotransmitters GABA and glutamate, the amino acid tryptophan, and short-chain fatty acids involved in myelination, including acetate and butyrate. Microbial gene sets around 4 months of age were strongly associated with the VEP from around 9 to 14 months of age and showed more associations than concurrently measured gene sets, suggesting microbial metabolism in early life may affect subsequent neural plasticity and development.

## Introduction

The gut microbiome in early life has potential long-term implications for brain and body health. One important way this influence can occur is through interactions with the central nervous system as a “microbial-gut-brain axis” (1–3). The metabolic potential of the microorganisms that inhabit the gut vastly exceeds that of human cells alone, with microbial genes outnumbering host genes by a hundredfold (4). In particular, gut microbes have the ability to metabolize and synthesize many neuroactive compounds (5). However, the physiological relevance of this in humans has been difficult to quantify, particularly during initial neurological development in early life.

Extensive work in preclinical models suggests that these neuroactive compounds can influence the brain through both direct and indirect pathways. For example, major neurotransmitters (e.g., glutamate, γ-aminobutyric acid (GABA), serotonin, and dopamine) are readily synthesized and degraded by intestinal microbes and can enter circulation and pass the blood-brain barrier to influence central nervous function (6–9). Glutamatergic/GABA-ergic signaling is critical for balancing the brain’s excitatory and inhibitory neurotransmission levels, and alterations in the bi-directional glutamatergic/GABA-ergic signaling between the gut microbiome and brain are implicated in several physical and mental health conditions (10,11). Similarly, the gut and the microbiome are critical to the regulation of metabolism for the neurotransmitters serotonin and dopamine, particularly through the metabolism of dietary tryptophan (12). Moreover, short-chain fatty acids (SCFAs) produced by the gut microbiome may impact the brain directly by modulating neurotrophic factors, glial and microglial maturation and myelination, and neuroinflammation (13,14). Other indirect pathways for gut microbial influence on the brain include vagus nerve stimulation, neuroendocrine modulation, and immune system regulation (1).

Rapidly growing literature connects the metabolic potential of the gut microbiome and brain function in humans (reviewed in 7,15,16), but the overwhelming majority of this research is conducted in adult participants. Importantly, both the gut microbiome and the brain undergo dramatic and rapid development over the first postnatal years (17–19). However, very little is currently known about how gut-brain influences emerge or change during this critical window (20–22). Interrogating this early co-development in humans is key to both understanding adaptive gut-brain function and behavior and informing strategies to support it. Specifically, the visual cortex has been shown to be sensitive to gut microbiome modulations in adults (23) and in rodents (24), yet the visual cortex undergoes its most rapid period of plasticity and maturation over infancy at the same time the microbiome changes most significantly (25–27). Visual cortical maturation can be robustly indexed via electroencephalography (EEG) with the Visual-Evoked Potential (VEP) response to visual stimuli from birth. The VEP morphology includes amplitude deflections, sensitive to neurotransmission changes, as well as latencies to those deflections, sensitive to structural changes. Moreover, the VEP is an important paradigm for indexing neurodevelopment given its translational potential, as it can be studied mechanistically across species and has clinical utility (28,29).

Here, we investigated the longitudinal co-development of microbial metabolic potential quantified via genes encoding enzymes that metabolize neuroactive compounds and visual neurodevelopment as indexed by the VEP in a longitudinal community sample of 194 infants from Gugulethu in Cape Town, South Africa, recruited as part of the prospective longitudinal “Khula” Study (30). Stool samples and EEG were each collected at up to 3 visits in the first 18 months of life. Shotgun metagenomic sequencing was used to obtain microbial gene sequences from infant stool samples. To index visual cortical functional development, latencies and peak amplitudes were extracted from each component of the VEP (i.e., first negative-going deflection, N1; first positive-going deflection, P1; and second negative-going deflection, N2), producing six VEP features of interest. We evaluated the concurrent association between microbial genes and VEP amplitudes and latencies, and we tested prospective influences of microbial genes from early visits on VEP changes at later visits. In this way, we were able to reveal the temporal dynamics of gut-brain co-development within individuals during this most critical window of plasticity in both systems.

## Methods and Materials

### Cohort

#### Participants and Study Design

Infants were recruited from local community clinics in Gugulethu, an informal settlement in Cape Town, South Africa, as part of a prospective longitudinal study (most enrollments happened prenatally with 16% of infants enrolled shortly after birth; 30). The first language for the majority of residents in this area is Xhosa. Study procedures were offered in English or Xhosa depending on the language preference of the mother. This study was approved by the relevant university Health Research Ethics Committees (University of Cape Town study number: 666/2021). Informed consent was collected from mothers on behalf of themselves and their infants. Demographic information, including maternal place of birth, primary spoken language, maternal age at enrollment, maternal educational attainment, and maternal income, were collected at enrollment (see Table 1).

**Table 1:** Overall Demographic Information.

|  | Overall<br>(N=194) |
| --- | --- |
| **Mean (SD) Age at EEG Data Collection (months)** |  |
| Visit 1 (N=97) | 3.7 (0.85) |
| Visit 2 (N=129) | 8.6 (1.46) |
| Visit 3 (N=130) | 14.1 (1.03) |
| **Mean (SD) Age at Stool Data Collection (months)** |  |
| Visit 1 (N=119) | 3.6 (0.76) |
| Visit 2 (N=105) | 8.8 (1.43) |
| Visit 3 (N=91) | 14.0 (1.24) |
| **Maternal Place of Birth** |  |
| South Africa | 191 (98.5%) |
| In the African Continent (not South Africa) | 3 (1.5%) |
| **Primary Spoken Language** |  |
| Xhosa Language | 187 (96.4%) |
| Sotho Language | 2 (1.0%) |
| Zulu Language | 1 (0.5%) |
| English Language | 2 (1.0%) |
| Ndebele Language | 1 (0.5%) |
| Afrikaans Language | 1 (0.5%) |
| **Maternal Age at Infant Birth (years)** |  |
| Mean (SD) | 29.2 (5.63) |
| Median [Min, Max] | 29.0 [18.0, 41.0] |
| Missing | 1 (0.5%) |
| **Maternal Educational Attainment<sup>a</sup>** |  |
| Completed Grade 6 (Standard 4) to Grade 7 (Standard 5) | 4 (2.1%) |
| Completed Grade 8 (Standard 6) to Grade 11 (Standard 9) i.e., high school without matriculating | 78 (40.2%) |
| Completed Grade 12 (Standard 10) i.e., high school | 88 (45.4%) |
| Part of university/ college/ post-matric education | 13 (6.7%) |
| Completed university/ college/ post-matric education | 11 (5.7%) |
| **Maternal Monthly Income<sup>b</sup> (South African Rand/ZAR)** |  |
| Less than R1000 per month | 97 (50.0%) |
| R1000 - R5000 per month | 76 (39.2%) |
| R5000 - R10,000 per month | 16 (8.2%) |
| More than R10,000 per month | 0 (0%) |
| Unknown | 5 (2.6%) |
| **Infant Biological Sex** |  |
| Female | 91 (46.9%) |
| Male | 103 (53.1%) |

<sup>a</sup>The South African Educational System was formerly divided into years called standards, similarly to the way the United States Educational System is divided into grades. The equivalent in terms of standards is provided in parentheses next to each mentioned grade. "University/College/Post-Matric Education" refers to tertiary or post-secondary education as defined by the World Bank.
<sup>b</sup>At the time of writing (1/16/24), 1 US Dollar = 18.87 South African Rand (ZAR).

Families were invited to participate in three in-lab study visits over their infant’s first two years of life. At the first in-lab study visit (hereafter visit-1), occurring when infants were between approximately 2 months and 6 months of age, the following data were collected: the infants’ age (in months), sex, infant electroencephalography (EEG), and infant stool samples.

At the second study visit (hereafter visit-2), occurring when infants were between approximately 6 months and 12 months of age (age in months: M=8.60, SD=1.48, range=5.41-12.00) and at the third study visit (hereafter visit-3), occurring when infants were between approximately 12 months and 17 months of age (age in months: M=14.10, SD=1.04, range=12.10-17.00), infant EEG and stool samples were collected again. At visits in which infants were unable to complete both EEG and stool samples on the same day, EEG and stool samples were collected on different days. For concurrent time point analyses, infants with EEG and stool collected more than two months apart were excluded. Not all infants had EEG and microbiome data collected at all three time points or contributed usable data at all three-time points.

All enrolled infants received a comprehensive medical exam at each visit, which included assessments of eye-related conditions. Several infants (n=3) were identified as having eye-related anomalies during the medical exam, and they were excluded from any further analyses.

### EEG Processing

#### EEG Data Acquisition

Electroencephalography (EEG) data were acquired from infants while they were seated in their caregiver’s lap in a dimly-lit, quiet room using a 128-channel high density HydroCel Geodesic Sensor Net (EGI, Eugene, OR), amplified with a NetAmps 400 high-input amplifier, and recorded via an Electrical Geodesics, Inc. (EGI, Eugene, OR) system with a 1000 Hz sampling rate. EEG data were online referenced to the vertex (channel Cz) through the EGI Netstation software. Impedances were kept below 100KΩ in accordance with the impedance capabilities of the high-impedance amplifiers. Geodesic Sensor Nets with modified tall pedestals designed for improving inclusion of infants with thick/curly/tall hair were used as needed across participants (31). Shea moisture leave-in castor oil conditioner was applied to hair across the scalp prior to net placement to improve both impedances and participant comfort (31). This leave-in conditioner contains insulating ingredients, so there is no risk of electrical bridging, and has not been found to disrupt the EEG signal during testing (unpublished data). Conditioning hair in this way allows for nets to lay closer to the scalp for curly/coily hair types and makes for more comfortable net removal at the end of testing.

The Visual-Evoked Potential (VEP) task was presented using Eprime 3.0 software (Psychology Software Tools, Pittsburgh, PA) on a Lenovo desktop computer with an external monitor 19.5 inches on the diagonal facing the infant (with monitor approximately 65 cm away from the infant). A standard phase-reversal VEP was induced with a black and white checkerboard (1cm x 1 cm squares within the board) stimulus that alternated presentation (black squares became white, white squares became black) every 500 milliseconds for a total of 100 trials. Participant looking was monitored by video and by an assistant throughout data collection. If the participant looked away during the VEP task, the task was rerun.

#### EEG Data Pre-Processing

VEP data were exported from native Netstation `.mff` format to `.raw` format and then pre-processed using the HAPPE+ER pipeline within the HAPPE v3.3 software, an automated open-source EEG processing software validated for infant data (32). A subset of the 128 channels were selected for pre-processing that excluded the rim electrodes as these are typically artifact-laden (channels excluded from pre-processing included in Table S4). The HAPPE pre-processing pipeline was run with user-selected specifications outlined in Table S4.

Pre-processed VEP data were considered usable and moved forward to VEP extraction if HAPPE pre-processing ran successfully, at least 15 trials were retained following bad trial rejection, and at least one good channel was kept within the visual ROI. Note that channels marked bad during pre-processing had their data interpolated as part of standard preprocessing pipelines for ERPs (32). Interpolated channels were included in analyses here as is typically done in developmental samples, and given the low overall rates of interpolation present (e.g., an average of between 4 to 5 of 5 possible good channels in the region of interest were retained at each visit time point).

#### Visual-Evoked Potentials (VEPs)

VEP waveforms were extracted and quantified using the HAPPE+ER v3.3 GenerateERPs script (32). Electrodes in the occipital region were selected as a region of interest (i.e., E70, E71, E75, E76, E83). The VEP waveform has three main components to be quantified: a negative N1 peak, a positive P1 peak, and a negative N2 peak. Due to normative maturation of the waveforms as infants age, one set of user-specified windows for calculating component features was used for visit-1 and 2 and another was used for visit-3. For visits 1 and 2, the window for calculating features for the N1 component was 40-100 ms, 75-175 ms for the P1 component, and 100-325 ms for the N2 component. For visit-3, the window for calculating features for the N1 component was 35-80 ms, 75-130 ms for the P1 component, and 100-275 ms for the N2 component. HAPPE+ER parameters used in extracting the ERPs are summarized in Table S5.

To correct for the potential influence of earlier components on later components, corrected amplitudes and latencies were calculated and used in all analyses. Specifically, the P1 amplitude was corrected for the N1 amplitude (corrected P1 amplitude = P1 -N1 amplitude), the P1 latency was corrected for the N1 latency (corrected P1 latency = P1 - N1 latency), the N2 amplitude was corrected for the P1 amplitude (corrected N2 amplitude = N2 - P1 amplitude), and the N2 latency was corrected for the P1 latency (corrected N2 latency = N2 - P1 latency).

All VEPs were visually inspected to ensure that the automatically extracted values were correct and were adjusted if observable peaks occurred outside the automated window bounds. Participants were considered to have failed this visual inspection and were subsequently removed from the data set if their VEP did not produce three discernible peaks. VEP waveforms of included participants by time point are included in Figure 2A. Ninety seven infants provided usable VEP data at visit-1, 130 infants provided usable VEP data at visit-2, and 131 infants provided usable VEP data at visit-3. For included participants, EEG data quality metrics are summarized in Table S6. T-tests for data quality metrics (i.e., number of trials collected, number of trials retained, number of channels retained in the ROI, Pearson’s r for data pre-vs. post-wavelet thresholding at 5, 8, 12, and 20 Hz) were run between each visit combination (i.e., visit-1 vs. visit-2, visit-1 vs. visit-3, visit-2 vs. visit-3). For visits that differed in data quality, follow-up post hoc correlations were run for the data quality measure with each VEP feature at each visit in the T-test. In no case did the data quality metric relate to VEP features at multiple visits, making it highly unlikely the data quality difference contributed to results.

### Biospecimens and sequencing

#### Sample Collection

Stool samples (n=315) were collected in the clinic by the research assistant directly from the diaper, transferred to Zymo DNA/RNA ShieldTM Fecal collection Tubes (#R1101, Zymo Research Corp., Irvine, USA) and immediately frozen at −80 °C. Stool samples were not collected if the participant had taken antibiotics within the two weeks prior to sampling.

#### DNA Extraction

DNA extraction was performed at Medical Microbiology, University of Cape Town, South Africa, from stool samples collected in DNA/RNA Shield™ Fecal collection tube using the Zymo Research Fecal DNA MiniPrep kit (# D4300, Zymo Research Corp., Irvine, USA) following manufacturer’s protocol. To assess the extraction process’s quality, ZymoBIOMICS® Microbial Community Standards (#D6300 and #D6310, Zymo Research Corp., Irvine, USA) were incorporated and subjected to the identical process as the stool samples. The DNA yield and purity were determined using the NanoDrop® ND −1000 (Nanodrop Technologies Inc. Wilmington, USA).

#### Sequencing

Shotgun metagenomic sequencing was performed on all samples at the Integrated Microbiome Research Resource (IMR, Dalhousie University, NS, Canada). A pooled library (max 96 samples per run) was prepared using the Illumina Nextera Flex Kit for MiSeq and NextSeq from 1 ng of each sample. Samples were then pooled onto a plate and sequenced on the Illumina NextSeq 2000 platform using 150+150 bp paired-end P3 cells, generating 24M million raw reads and 3.6 Gb of sequence per sample (33).

### Statistics / computational analysis

#### Age-Related Changes in VEP Features

To determine age-related changes in VEP features, six linear mixed models with each VEP feature as the outcome (i.e., N1 amplitude/latency, P1 amplitude/latency, N2 amplitude/latency) were run using the lme4 package (34) in R with age in months as the predictor of interest and number of retained trials as a covariate.

#### Metagenome processing

Raw metagenomic sequence reads (2.5 x 10⁷ ± 1.4 x 10⁷ reads/sample) were processed using tools from the bioBakery as previously described (17,35). Briefly, KneadData v0.10.0 was used with default parameters to trim low-quality reads and remove human sequences (using reference database hg37). Next, MetaPhlAn v3.1.0 (using database mpa_v31_CHOCOPhlAn_201901) was used with default parameters to map microbial marker genes to generate taxonomic profiles. Taxonomic profiles and raw reads were passed to HUMAnN v3.7 to generate stratified functional profiles.

#### Microbial community analysis

Principal coordinates analysis was performed in the julia programming language (36) using the Microbiome.jl package (37). Bray-Curtis dissimilarity (Distances.jl) was calculated across all pairs of samples, filtering for species-level classification. Classical multidimensional scaling was performed on the dissimilarity matrix (MultivariateStats.jl), and axes with negative eigenvalues were discarded.

#### Feature Set Enrichment Analysis (FSEA)

Potentially neuroactive genesets were extracted from Supplementary Dataset 1 from (5). Gut-brain modules provide Kegg Orthologue IDs (KOs) (38,39), which were mapped to UniRef90 IDs using the utility mapping file provided with HUMAnN v3.1 (35). For each stool/VEP pair, logistic regression (LR) was performed, linking the presence or absence of that UniRef in a sample with each VEP feature (i.e., N1, P1, and N2 latencies and amplitudes), controlling for the age at which the stool sample was collected, the number of retained VEP trials, and the difference in age between the stool collection and VEP measurement. For concurrently collected stool and VEP comparisons (Figure 2, Figure S2), participants whose stool collection and VEP measurements were more than two months apart were excluded.

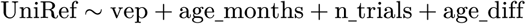

FSEA was performed on each geneset that had at least five members in each comparison group according to the procedure set out in Subramanian *et. al.* (2005) (40). Briefly, enrichment scores (ES) are calculated based on the rank order of z-statistics from the LR for each UniRef. A permutation test was then performed where the ES for 5000 random samples of ranks of the same length as the gene set are calculated, and the pseudo-p value is the fraction of permutations where the permutation ES has a greater absolute value than the true ES.

Benjamini-Hochberg FDR correction was performed separately on all concurrently tested geneset/ VEP feature combinations and all longitudinal geneset/VEP feature combinations. Corrected p-values (q-values) less than 0.2 were considered statistically significant.

For longitudinal comparisons, all participants that had a stool sample collected at one visit and a VEP assessment at a subsequent visit were included (visit-1 stool → visit-2 VEP, N = 84; v1 → v2, N = 76; v2 → v3, N = 69). A total of 95 geneset/VEP features were significant when using an FDR-corrected p-value cutoff of q < 0.2. To ensure the robustness of these findings, we randomly permuted participant IDs between the stool and VEP assessments and repeated the analysis. Over 10 random permutations, a mean of 19.5 significant associations were identified, suggesting that FDR correction is correctly calibrating the false-positive rate.

#### Data and code availability

Code for initial processing of data and for analyses performed in this manuscript are available on github and archived on Zenodo (41). Input data will be archived on Dryad and downloadable via included scripts in analysis code upon publication.

## Results

### The brain and microbiome develop rapidly in the first months of life

To investigate the co-development of the gut microbiome and visual neurodevelopment, we collected stool and the VEP in a longitudinal cohort of 194 children in South Africa during the first 18 months of life (Figure 1A, B, Table 1; visit-1, N = 119, age 3.6 ± 0.7 months, visit-2, N = 144, age 8.7 ± 1.4 months, visit-3, N = 130, age 14.2 ± 1.0 months). As expected for children at this age, both amplitude and latency VEP features were strongly correlated with age (18,42), That is, as infants got older, N1 amplitude became more negative (N1: b=-0.07, p<.05), corrected P1 amplitude became smaller (P1: b=-0.50, p<.05), corrected N2 amplitude became smaller (N2: b=0.52, p<.05), and all latencies became shorter (N1: b=-0.79, p<.05; P1: b=-1.21, p<.05; N2: b=-4.10, p<.05) (Figure 2A).

**Figure 1:**
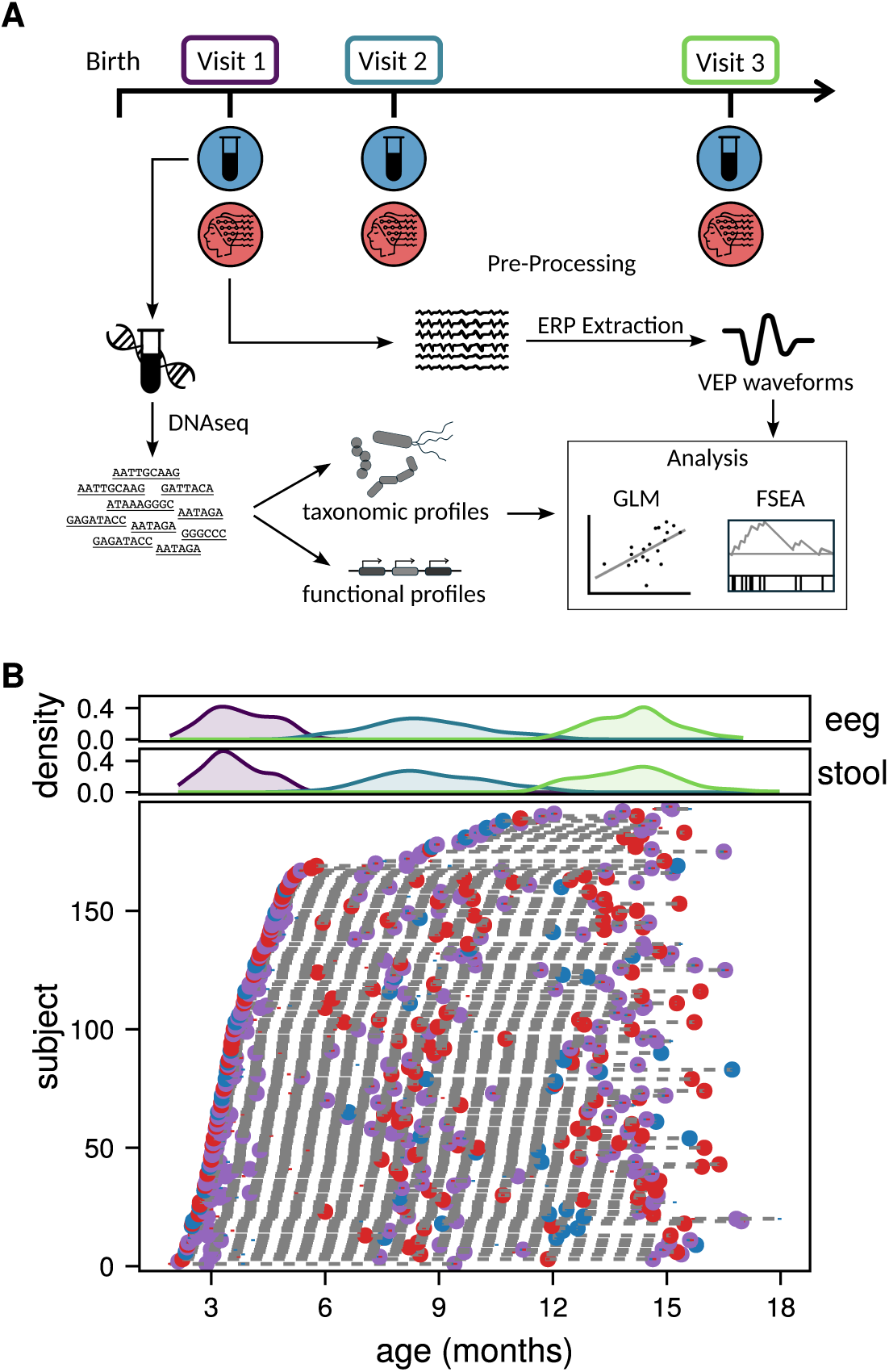
Study design to capture the dynamic nature of early microbiome and brain development. (A) Study design; participants (N=194) were seen up to 3 times over the first 18 months of life. Stool samples and EEG data were collected, generating microbial functional profiles (stool) and VEP waveforms (EEG) used in subsequent analyses. (B) Longitudinal sampling of study participants; Density plots (top) for stool and EEG collection show the ages represented in each visit. The scatter plot (bottom) shows individual participant visits. Dotted lines connect separate visits for the same participant. When stool and EEG data were collected for the same visit (purple) but not on the same day, dot represents the median age of collection, and vertical bars in blue and red represent stool and EEG collections, respectively.

Similarly, microbial composition was developmentally dependent, as expected (42–44). Ordinations reveal a similar relationship with age, with the first principal coordinate axis for both taxonomic profiles (Figure 2C; variance explained = 15.1%; R = −0.50) and functional profiles (Figure 2D; variance explained = 12.9%; R = −0.57) driven strongly by the age of the participant at the time of collection. Early samples were dominated by *Bifidobacterium* and *Bacteroides* species, while later samples have increasing *Prevotella* and anaerobic genera such as *Faecalibacterium* (Figure 2E).

**Figure 2:**
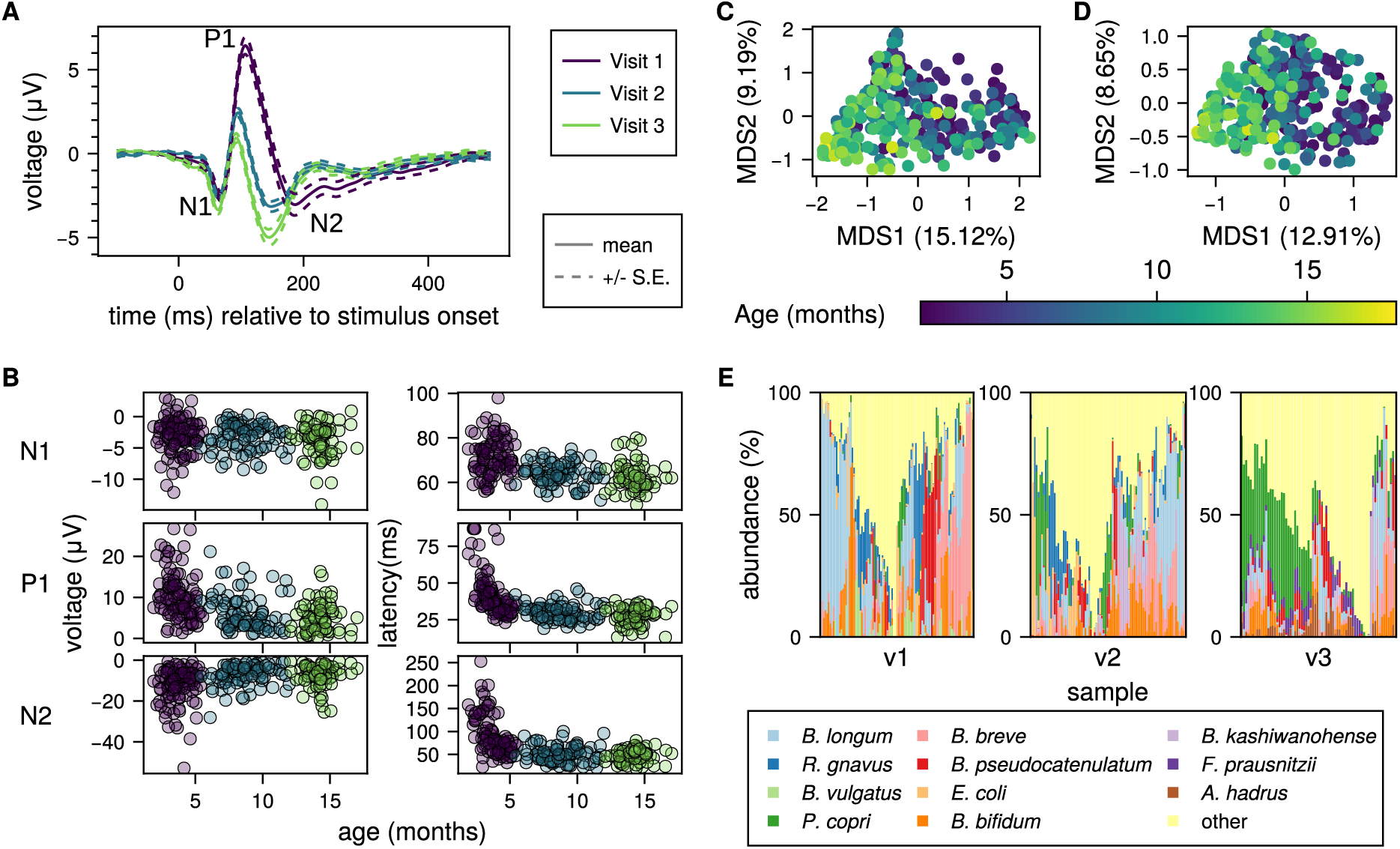
The gut microbiome and VEP both develop over the first 18 months of life. (A) Mean ± standard error of VEP curves from all included individuals at each visit. (B) Individual VEP feature measurements for peak amplitudes (left) and latencies (right) for all participants and all visits in the study, separated by age and colored by visit as in (A). (C) Principal coordinate analysis (PCoA) by multidimensional scaling (MDS) on Bray-Curtis dissimilarity of taxonomic profiles; percent variance explained (fraction of positive eigenvalues) by each of the first two axes are indicated on the x and y axes respectively. (D) PCoA of microbial functional profiles (UniRef90s). (E) Relative abundance of the top 11 microbial species across all visits. All other species were summed so that the total abundance is 100%. Each column represents a single sample, and samples are ordered by hierarchical clustering based on Bray-Curtis dissimilarity of the full microbial composition.

### Microbial genes with neuroactive potential are associated with concurrently measured visual development

To test whether microbial metabolic potential was related to early life brain activity, we performed feature set enrichment analysis (FSEA) using previously-defined groups of potentially neuroactive microbial genes and the concurrently measured VEP amplitude and latency features (5,17). For each gene set that had at least 5 genes represented in a given comparison group, logistic regression was performed using VEP features as predictors and the presence or absence of each microbial gene in the metagenome as the response to determine concurrent associations (see Methods). Z statistics for in-set genes were compared to all genes using a permutation test to determine significance of the associations (40).

Of the 35 genesets assessed, 19 had sufficient representation to test, and of those, 18 were significantly associated with at least one EEG feature during at least one visit within the 18-month window, after correcting for false discovery rate (Benjamini-Hochberg, q < 0.2; Figure 3, Table S2). Microbial genes involved in synthesis or degradation of molecules with neuroactive potential across all categories considered (i.e., neurotransmitters, amino acid metabolism, SCFAs, other) were associated with both concurrent VEP amplitudes and latencies at each visit (Figure S2), demonstrating widespread associations between early life gut microbiome and visual cortex neurodevelopment. The number of these concurrent associations increased over time (visit-1 had 6 associations, visit-2 had 24, and visit-3 had 37).

**Figure 3:**
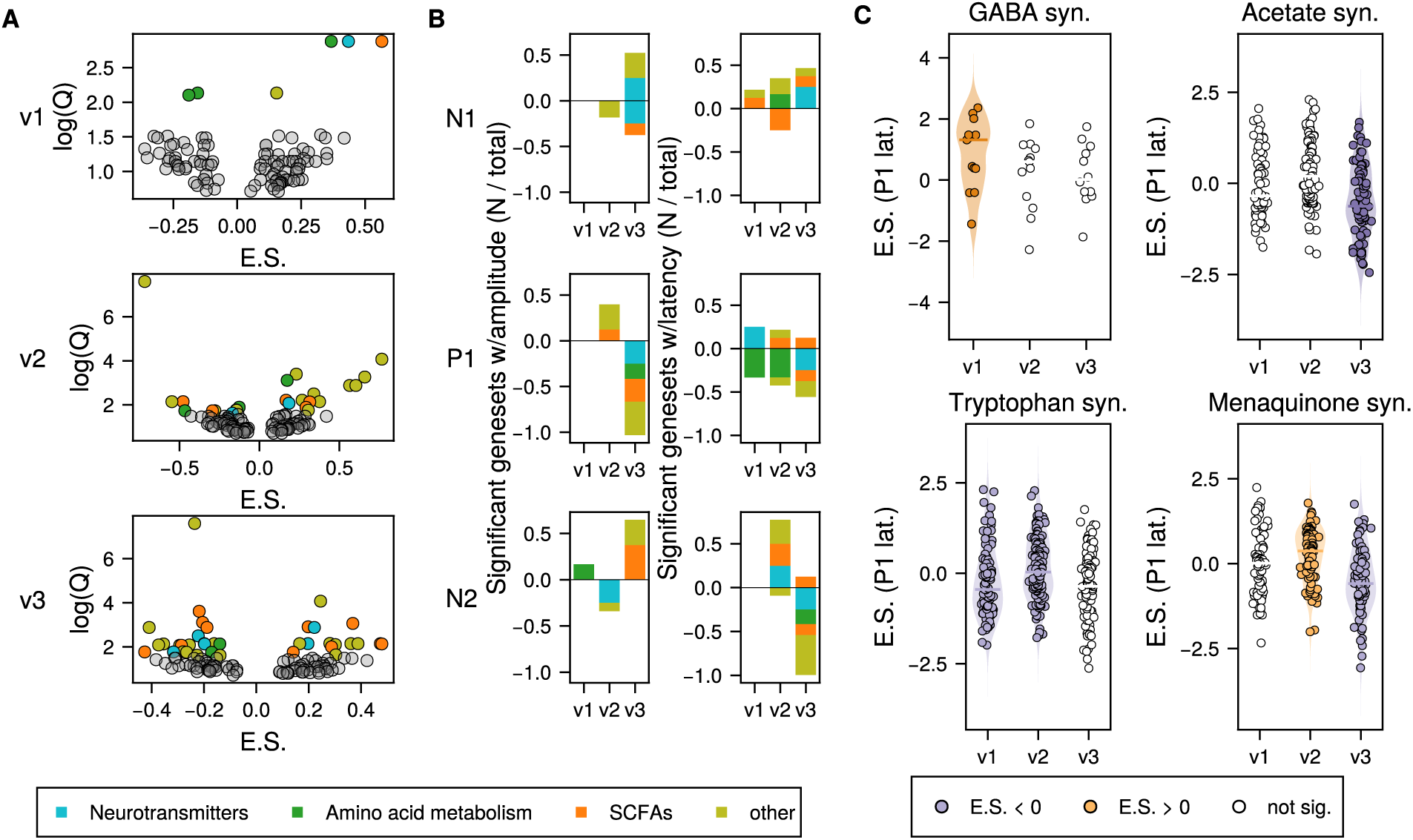
Feature Set Enrichment Analysis Reveals associations between microbial genes and VEP. (A) Volcano plots of of gene sets tested with feature set enrichment analysis (FSEA) for all 6 VEP features for each visit (visit-1 top, visit-2 middle, visit-3 bottom) with enrichment score (E.S.) compared to log scaled FDR-corrected p-value (Q). Colored dots were significantly enriched (positive E.S.) or depleted (negative E.S.) relative to the tested VEP feature. (B) Summary of results in (A), showing the fraction of each class of neuroactive genes (neurotransmitter metabolism, SCFA metabolism, amino acid metabolism, or other) that were statistically significantly enriched or depleted for each VEP feature for each visit in the analysis. (C) Enrichment plots for selected gene sets and their association with P1 latency. Each plot shows the distribution of associations of individual genes within the gene set and the VEP feature. Dots are colored if the geneset as a whole was significantly associated. Enrichment plots for all gene set / VEP feature associations are shown in Figure S2.

Specifically, across the gene sets involved in neurotransmitter synthesis and degradation, glutamate synthesis/degradation and GABA synthesis showed associations with all VEP features, primarily at the 2nd and 3rd visit (mean ages 8.6 and 14.1 months, respectively; Figure 3C Table S2). Gene sets involved in tryptophan metabolism and associated pathways (i.e., quinolinic acid) were also strongly concurrently related to VEP development.

Several short-chain fatty acid (SCFA)-metabolizing gene sets were also found to have multiple associations with VEP features. Specifically, acetate synthesis was strongly associated with almost all VEP features (Table S2). Butyrate synthesis was associated with P1 and N2 amplitudes and latencies from 6-months onwards (visits 2 and 3), when the visual cortex is most actively undergoing myelination. Lastly, propionate synthesis/degradation was significantly associated with VEP latencies at every visit over the 18-month window (N1 at visits 1 and 2, and both P1 and N2 at visit-3). These SCFA metabolizing genes showed almost double the concurrent associations with VEP latencies than amplitudes (11 associations with latencies, 6 associations with amplitudes).

Finally, within the remaining gene sets tested, we observed robust associations in particular between menaquinone (Vitamin K2) gene sets and the VEP features over this infancy window. This is an expected relationship, as vitamin K2 specifically is posited to promote healthy vision, both outside of the brain through effects on the retina, and within the brain where it can protect neural circuits from oxidative stress (45).

Notably, across significant gene set associations with VEP features, the P1 and N2 component amplitudes and latencies were consistently the most sensitive to these microbial gene sets. Both P1 and N2 components are known to show the most protracted and dramatic changes with development during the first year of life (46) and may best reflect underlying visual learning and plasticity at this stage (Table S1).

### Microbial metabolic potential predicts future brain development in infancy

We initially hypothesized that the earliest microbial influences would have the largest effects on brain development, but in cross-sectional analysis with concurrently measured VEP, we observed the fewest number of associations at visit-1. To differentiate whether this cross-sectional finding indicated the early microbiome was sparsely related to visual cortical development or instead took time to manifest its influence, we sought to determine whether microbial genes at early time points were associated with later VEP development. We therefore performed FSEA on stool samples collected at visit-1 with visit-2 VEP (see Table 2, age at stool collection = 3.6 ± 0.8 months, age at VEP = 8.6 ± 1.5 months) or visit-3 VEP (see Table 3, age at stool collection = 3.7 ± 0.7 months, age at VEP = 14.1 ± 1.1 months), as well as visit-2 stool samples with visit-3 VEP (see Table 4, age at stool collection = 8.9 ± 1.5 months, age at VEP = 14.3 ± 1.0 months; Table S3) (Figure 4A, Figure S3).

**Table 2:**
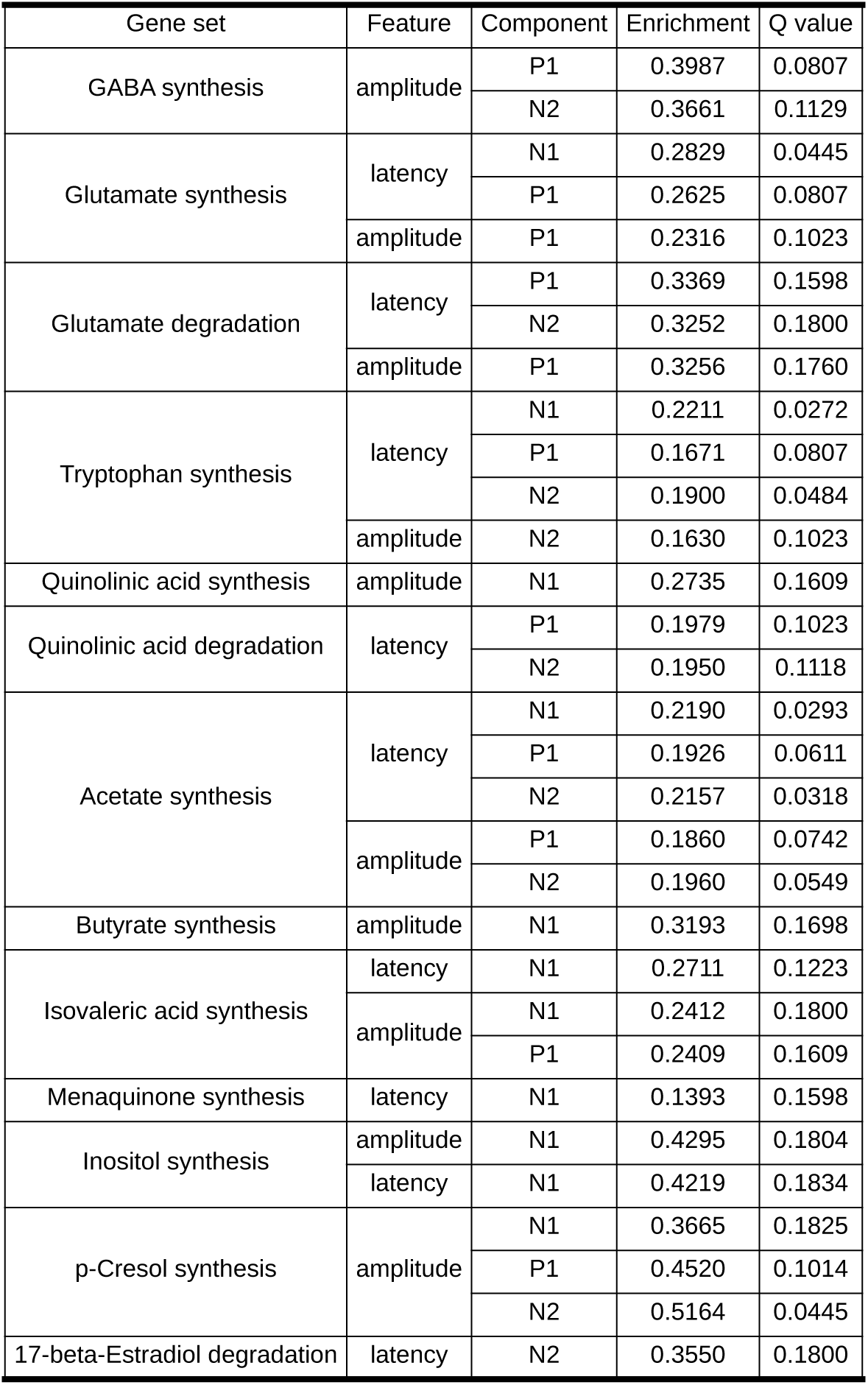
Longitudinal FSEA, visit-1 stool ⇨ visit-2 VEP.

| Gene set | Feature | Component | Enrichment | Q value |
| --- | --- | --- | --- | --- |
| GABA synthesis | amplitude | P1 | 0.3987 | 0.0807 |
|  |  | N2 | 0.3661 | 0.1129 |
| Glutamate synthesis | latency | N1 | 0.2829 | 0.0445 |
|  |  | P1 | 0.2625 | 0.0807 |
|  | amplitude | P1 | 0.2316 | 0.1023 |
| Glutamate degradation | latency | P1 | 0.3369 | 0.1598 |
|  |  | N2 | 0.3252 | 0.1800 |
|  | amplitude | P1 | 0.3256 | 0.1760 |
| Tryptophan synthesis | latency | N1 | 0.2211 | 0.0272 |
|  |  | P1 | 0.1671 | 0.0807 |
|  |  | N2 | 0.1900 | 0.0484 |
|  | amplitude | N2 | 0.1630 | 0.1023 |
| Quinolinic acid synthesis | amplitude | N1 | 0.2735 | 0.1609 |
| Quinolinic acid degradation | latency | P1 | 0.1979 | 0.1023 |
|  |  | N2 | 0.1950 | 0.1118 |
| Acetate synthesis | latency | N1 | 0.2190 | 0.0293 |
|  |  | P1 | 0.1926 | 0.0611 |
|  |  | N2 | 0.2157 | 0.0318 |
|  | amplitude | P1 | 0.1860 | 0.0742 |
|  |  | N2 | 0.1960 | 0.0549 |
| Butyrate synthesis | amplitude | N1 | 0.3193 | 0.1698 |
| Isovaleric acid synthesis | latency | N1 | 0.2711 | 0.1223 |
|  | amplitude | N1 | 0.2412 | 0.1800 |
|  |  | P1 | 0.2409 | 0.1609 |
| Menaquinone synthesis | latency | N1 | 0.1393 | 0.1598 |
| Inositol synthesis | amplitude | N1 | 0.4295 | 0.1804 |
|  | latency | N1 | 0.4219 | 0.1834 |
| p-Cresol synthesis | amplitude | N1 | 0.3665 | 0.1825 |
|  |  | P1 | 0.4520 | 0.1014 |
|  |  | N2 | 0.5164 | 0.0445 |
| 17-beta-Estradiol degradation | latency | N2 | 0.3550 | 0.1800 |

**Table 3:** Longitudinal FSEA, visit-1 stool ⇨ visit-3 VEP.

| Gene set | Feature | Component | Enrichment | Q value |
| --- | --- | --- | --- | --- |
| GABA synthesis | latency | P1 | 0.4343 | 0.0445 |
|  | amplitude | P1 | 0.4099 | 0.0742 |
|  |  | N2 | 0.5065 | 0.0254 |
| Glutamate synthesis | latency | N1 | 0.3035 | 0.0293 |
|  | amplitude | N1 | 0.2344 | 0.1118 |
| Tryptophan synthesis | latency | N1 | 0.1875 | 0.0445 |
|  | amplitude | P1 | 0.2176 | 0.0159 |
| Quinolinic acid degradation | latency | P1 | 0.1862 | 0.1093 |
|  |  | N2 | 0.2624 | 0.1834 |
|  | amplitude | P1 | 0.1689 | 0.1609 |
| Acetate synthesis | latency | N1 | 0.1728 | 0.0807 |
| Propionate synthesis | amplitude | P1 | 0.3586 | 0.0807 |
| Propionate degradation | latency | N1 | 0.5097 | 0.1093 |
|  |  | P1 | 0.5345 | 0.0880 |
|  | amplitude | P1 | 0.7388 | 0 |
|  |  | N2 | 0.6669 | 0.0272 |
| Butyrate synthesis | latency | N1 | 0.3373 | 0.1223 |
|  |  | N2 | 0.3232 | 0.1498 |
|  | amplitude | P1 | 0.3688 | 0.1014 |
| Menaquinone synthesis | latency | P1 | 0.1582 | 0.0870 |
|  | amplitude | N1 | 0.1417 | 0.1397 |
|  |  | P1 | 0.1488 | 0.1131 |
|  |  | N2 | 0.1567 | 0.1023 |
| Inositol synthesis | amplitude | N2 | 0.4538 | 0.1210 |
| ClpB | latency | N1 | 0.2835 | 0.1804 |
|  | amplitude | P1 | 0.3284 | 0.1129 |
|  |  | N2 | 0.3585 | 0.0800 |

**Table 4:** Longitudinal FSEA, visit-2 stool ⇨ visit-3 VEP.

| Gene set | Feature | Component | Enrichment | Q value |
| --- | --- | --- | --- | --- |
| GABA synthesis | latency | N1 | 0.3762 | 0.1073 |
| Glutamate synthesis | latency | N1 | 0.1676 | 0.1609 |
|  |  | P1 | 0.2979 | 0.0159 |
|  | amplitude | P1 | 0.2342 | 0.0381 |
| Glutamate degradation | latency | P1 | 0.3174 | 0.1404 |
|  | amplitude | N2 | 0.2829 | 0.1875 |
| Tryptophan synthesis | amplitude | N1 | 0.1554 | 0.0608 |
|  |  | P1 | 0.1865 | 0.0293 |
| Quinolinic acid synthesis | latency | N1 | 0.3186 | 0.0610 |
|  |  | P1 | 0.3331 | 0.0437 |
|  | amplitude | P1 | 0.2577 | 0.1391 |
| Acetate synthesis | latency | N1 | 0.1673 | 0.0610 |
|  |  | P1 | 0.2574 | 0.000 |
|  |  | N2 | 0.1373 | 0.1529 |
|  | amplitude | P1 | 0.1469 | 0.1210 |
| Propionate synthesis | amplitude | P1 | 0.2994 | 0.1262 |
| Propionate degradation | latency | N1 | 0.5503 | 0.0742 |
|  |  | N2 | 0.4492 | 0.1609 |
|  | amplitude | P1 | 0.5088 | 0.1118 |
|  |  | N2 | 0.4229 | 0.1875 |
| Butyrate synthesis | latency | N1 | 0.3316 | 0.0807 |
|  |  | P1 | 0.3335 | 0.0742 |
|  |  | N2 | 0.2908 | 0.1223 |
|  | amplitude | N2 | 0.3066 | 0.1118 |
| Isovaleric acid synthesis | latency | P1 | 0.2316 | 0.1556 |
|  | amplitude | P1 | 0.2469 | 0.1129 |
| Menaquinone synthesis | latency | N1 | 0.1528 | 0.1210 |
|  | amplitude | N1 | 0.1644 | 0.1073 |
|  |  | P1 | 0.2014 | 0.0293 |
|  |  | N2 | 0.1634 | 0.1121 |
| Inositol degradation | latency | N1 | 0.6381 | 0.0293 |
|  |  | P1 | 0.5323 | 0.1014 |
|  | amplitude | P1 | 0.6413 | 0.0293 |
| p-Cresol synthesis | latency | P1 | 0.3118 | 0.1397 |
|  |  | N2 | 0.3152 | 0.1391 |
|  | amplitude | N1 | 0.3039 | 0.1658 |
| S-Adenosylmethionine synthesis | amplitude | N1 | 0.2228 | 0.1556 |
|  |  | P1 | 0.2295 | 0.1347 |
| 17-beta-Estradiol degradation | amplitude | P1 | 0.2818 | 0.1875 |
| ClpB | latency | P1 | 0.2687 | 0.1804 |

**Figure 4:**
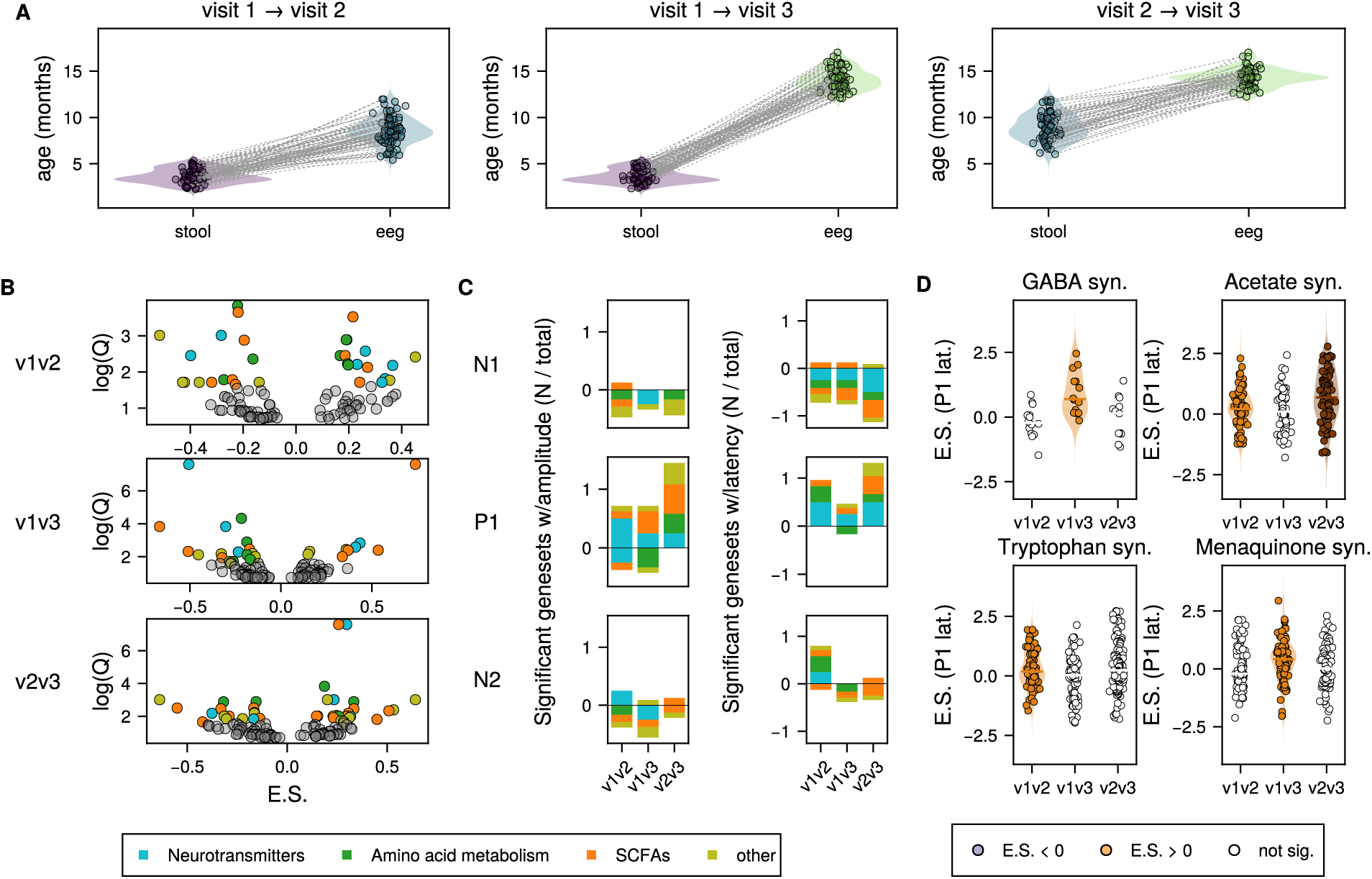
(A) Age distributions for cross-visit comparisons, with age at stool collection (left) and age of VEP measurement (right) for each participant included in the analysis. Collections for the same individual are connected by a dotted gray line. (B) Volcano plots of of gene sets tested with feature set enrichment analysis (FSEA) for all 6 VEP features, with enrichment score (E.S.) compared to log scaled FDR-corrected p-value (Q). Colored dots were significantly enriched (positive E.S.) or depleted (negative E.S.) relative to the tested VEP feature. (C) Summary of results in (B), showing the fraction of each class of neuroactive genes (neurotransmitter metabolism, SCFA metabolism, amino acid metabolism, or other) that were statistically significantly enriched or depleted for each VEP feature for each cross-visit comparison in the analysis. (D) Enrichment plots for selected gene sets and there association with P1 latency. Each plot shows the distribution of associations of individual genes within the gene set and the VEP feature. Dots are colored if the geneset as a whole was significantly associated. Enrichment plots for all gene set / VEP feature associations may be found in Figure S2.

All gene sets that had a significant hit with concurrently measured VEP were also significantly associated with at least one future VEP featuree, except those involved in the synthesis of 3,4-dihydroxyphenylacetic acid (DOPAC), a metabolite of dopamine (Figure S3B, Tables 2-5). Notably, the quantity of those associations increased substantially for all longitudinal comparisons compared to concurrent comparisons. For example, only 6 visit-1 microbial gene sets were associated with visit-1 VEP, and each of those was only associated with a single concurrently measured VEP feature. By contrast, these longitudinal analyses revealed that visit-1 microbial gene sets showed a much richer pattern of associations with future VEP feature development. Specifically, 13 visit-1 gene sets were associated with visit-2 VEP features, and 11 were associated with visit-3 VEP features, the majority (9/13 for visit-2, 8/11 for visit-3) were associated with at least 2 VEP features, and nearly half (6/13 for visit-2, 5/11 for visit-3) were associated with more than 2 future VEP features.

Longitudinally, the early microbiome (visit-1) was related to VEP features at visit-2 and visit-3 fairly evenly (12/28 visit-1 microbiome associations to visit-3 VEP latencies, 15/30 associations to visit-3 amplitudes), suggesting early microbiome metabolism in the first 6 months of life is associated with visual neurodevelopment over the next year. Microbiome metabolism from visit-2 was associated with similar numbers of visit-3 VEP features as visit-1 microbiome (17 visit-3 latency features, 18 visit-3 amplitude features), suggesting continued co-development of these systems over the first postnatal year. Neurotransmitters GABA and glutamate, tryptophan metabolism (tryptophan and quinolinic acid), SCFAs including acetate, butyrate, and propionate, as well as menaquinone (Vitamin K2) were again all significantly associated with multiple VEP features across multiple longitudinal comparisons.

Importantly, the nature and identity of the longitudinal associations varied over development for many gene sets, indicating temporal specificity to these associations. With respect to the neurotransmitter-related pathways, amongst the associations between GABA synthesis genes and future VEP features, GABA genes specifically from visit-1 showed the greatest number of associations with future VEP features (5/6 GABA associations) at visits 2 and 3 equally, and the majority of these associations were VEP amplitudes (reflecting development of neurotransmission including excitatory/inhibitory balance). To a lesser extent, glutamate metabolism genes followed a similar temporal pattern (8/13 glutamate associations involved visit-1 genes) but did not relate to amplitudes or latencies differentially like GABA. This pattern of results suggests early (within the first 6 postnatal months) microbiome GABA/glutamate dynamics, especially GABA, are most relevant for changes to visual cortex function over the following year.

Tryptophan-related pathway genes (responsible for generating serotonin, amongst other products) from visit-1 were also responsible for the majority of associations with future VEP features (tryptophan: 6/8 associations; quinolinic acid: 6/9 associations). In contrast to GABA but similar to glutamate, tryptophan-related gene associations were largely shorter-term associations with VEP features at the visit immediately following gene set measurement (approximately 5 months later; 12/17 associations), indicating dynamic co-development over the first 18 months of life. Across neurotransmitter-related gene set associations (GABA, glutamate, tryptophan/serotonin), there was thus a clear pattern whereby early (∼4 months old) microbiome gene sets showed the largest number of associations with subsequent VEP feature development.

SCFAs showed a different developmental pattern of associations with future VEP features. Specifically, propionate and butyrate metabolism genes from both visit-1 and visit-2 showed associations with future VEP features, but here the effects were almost entirely observed for VEP features at visit-3 (10/10 propionate and 7/8 butyrate associations). Moreover, acetate and butyrate metabolism genes were doubly associated with future VEP latencies compared to amplitude features.

Finally, menaquinone (Vitamin K2) metabolism genes followed a similar pattern to the SCFAs in that genes from visit-1 and visit-2 were largely associated with future VEP features at visit-3 (8/9 menaquinone associations). This indicates persistent associations of this early microbiome gene set with individual differences in VEP features early in the second year of life.

It is possible that extrinsic factors related to development mutually influence both the gut microbiome and neural development, though we additionally tested whether VEP features were associated with microbial metabolism at a future visit, and found substantially fewer associations (29 total associations, compared with 95 when analyzing early stool samples with future VEP). While this does not prove a causal relationship, it is consistent with the hypothesis that microbial metabolism influences brain development.

## Discussion

The past decade has seen a remarkable growth in our understanding of the relations between the gut microbiome and the brain. However, a great deal of that investigation has focused on adult populations or neuropsychiatric disorders, limiting the potential to explain how and when these associations emerge during development. Here, we address this key open question by leveraging a rich longitudinal dataset over the first year and a half of life, which is the time of greatest developmental change for both the microbiome and brain given the unfolding of foundational sensory neurodevelopment. Our data revealed that microbial genes involved in the metabolism of neuroactive molecules are associated with concurrent and subsequent visual cortical neurodevelopment. These pathways included those for the neurotransmitters GABA and glutamate, the amino acid tryptophan, and short-chain fatty acids involved in myelination, including acetate and butyrate.

Specifically, we have shown a robust, prospective relationship between microbial genes involved in the metabolism of neuroactive compounds and the development of visual cortical function as measured by the VEP electrophysiological response. We found that microbial metabolism is more strongly associated with future measures of the VEP than those collected concurrently. While not dispositive, this would be the predicted outcome if microbial genes are causally influencing brain development. As additional evidence for this interpretation, we did not observe the same rich set of associations in the converse analyses examining whether VEP related to future microbiome properties. Microbial metabolism within the first 6 months shows the most associations with subsequent visual neurodevelopment, suggesting the early postnatal microbiome may play a particularly important role in the co-development of these systems. This interpretation is also supported by prior research showing that associations of the microbiome with behavioral readouts of neurocognition are stronger prospectively than concurrently (47). Moreover, specific associations between gene sets and VEP features showed temporal specificity within the 18-month developmental window assessed, suggesting that the impact of early microbial metabolism on the brain is developmentally dependent.

Notably, the gene sets most highly associated with visual functional neurodevelopment over infancy are for the metabolism of molecules with known links to developmental neuroplasticity (48–50). Specifically, we observed associations for gene sets related to glutamate and GABA, neurotransmitters that are central to regulating excitatory/inhibitory (E/I) cortical balance. Developmental changes in E/I balance modulate the degree of neuroplasticity in the mammalian cortex, including regulating the start and progression of critical period neuroplasticity mechanisms in the visual cortex (28,29,48,49). Our observed pattern of results suggests early (within the first 6 postnatal months) microbiome GABA/ glutamate dynamics, especially GABA, are most relevant for changes to visual cortex function over the following year. Gut production of GABA may influence cortical GABA levels via active transport from bloodstream to brain (51–53). Recent evidence suggests gut-derived glutamate may also influence brain levels and function (8–10) and can operate via indirect mechanisms (either transformation into GABA or via regulating glutamate levels in the bloodstream that impact glutamate transfer from brain to bloodstream).

Tryptophan related pathway genes were also identified here that are responsible for generating serotonin as well as other neuroactive molecules such as kynurenic acid (an SMDAR antagonist) (12). Both serotonin and kynurenic acid are implicated in early neuroplasticity and neurotransmitter regulation, and serotonin has potent effects for visual cortex plasticity in particular (54,55). While quinolinic acid is part of the kynurenine pathway and is a neurotoxin that can cause neuronal dysfunction, it may also play a role in glutamate uptake in the brain (56,57). Specifically, tryptophan metabolism genes were associated with VEP latencies just after each VEP component showed its greatest window of developmental change (components emerge sequentially as follows: P1, N1, N2). This pathway may thus relate to processes stabilizing the neural circuitry (i.e. downregulating neuroplasticity) underlying each VEP component, an account consistent with recently observed effects of serotonin within the visual cortex in rodents (58). Importantly, nearly all of the body’s serotonin is produced in the gut by enterochromaffin cells, and this biosynthesis is regulated by microbes (59,60), making this pathway an especially promising candidate intervention target for future research in development.

We further found that gene sets for short-chain fatty acids important in downregulating neuroinflammation and promoting myelination within the brain were robustly related to visual neurodevelopment. Myelination is important for down-regulating plasticity in neural circuitry over development by stabilizing and protecting circuits that have been shaped by early experience (61). Specifically, we observed associations between acetate, butyrate, and proprionate genes with VEP development. Acetate is a critical component required for the increased lipid synthesis that happens during postnatal myelination in the brain (62). Circulating butyrate also increases myelination (63), and though propionate’s relation to myelinating oligodendrocytes remains unclear, it is known to protect myelinating Schwann cells outside of the brain from oxidative stress (64). Acetate, butyrate, and propionate are all also widely regarded as neuroprotective by promoting healthy microglial development and downregulating neuroinflammation that interferes with myelination (65). VEP latency features reflect myelination (29,66,67), and accordingly, these SCFAs showed more associations with VEP latency features prospectively, especially VEP latency features in vsit 3. This pattern of results is consistent with these SCFA roles’ in myelination occurring over the second half of the developmental window studied. SCFAs including acetate, butyrate, and propionate can pass the blood-brain barrier to directly influence myelination-related processes within the brain. Taken together, the pattern of results across GABA, glutamate, tryptophan, and SCFA gene sets suggests early postnatal microbiome-derived metabolites relate to key neuroplasticity regulation processes within the cortex.

Our study is a substantial advancement over prior work on the microbial-gut-brain axis in early life due to the sequencing method, the large number of participants, the longitudinal study design, and the inclusion of participants from scientifically under-represented region of the world. The use of shotgun metagenomic sequencing enables direct interrogation of microbial metabolic potential. Prior research primarily used amplicon (16S rRNA gene) sequencing, which enables lower-resolution taxonomic identification and is restricted to inferring metabolic potential based on taxonomy. Moreover, several studies in infancy have inferred gut-brain associations by linking microbiome measures to subsequent neurodevelopmental measures using behavioral assessments (e.g., Bayley Scales of Infant Development), noting associations with visually-mediated cognition (47). However, this study assessed gut-brain associations directly using the VEP derived from electroencephalography. The VEP is advantageous because features reflect largely neurotransmission-related (via amplitudes) or structural (i.e., myelination, via latencies) changes over this developmental window, facilitating some specificity in the observed associations. Moreover, the VEP can be indexed with fidelity from birth, providing a continuous measure of visual cortical function across the study age-range. Additionally, this study involved a large number of participants (194) contributing dense longitudinal data, with up to three time points, all taken in the first 18 months of an infant’s life. While prior work focused on single time point measures of microbiome and neurodevelopment (68), longitudinal associations allowed us to investigate the changing relation between gut microbial metabolism and the development of visual neurocircuitry over time.

One limitation of this study is the fact that we are only able to observe the genomic composition of the microbiome, rather than the concentration of metabolites themselves. This prevents us from determining the concentration of these molecules in the gastrointestinal tract, blood, and brain, as the abundance of these genes does not provide information about their activity, their interactions with other metabolic pathways (including those of the host), or absorption by colonic epithelial cells. Moreover, the relationship between gene abundance and molecule concentration may be counterintuitive, since the relationship between degradation and synthesis of metabolites occurs both at the individual organism level and at the community level. For example, genes for breaking down a molecule may be prevalent if that molecule is at high concentrations, or the molecules may be rapidly degraded by other members of the community the moment they are produced. Furthermore, it may be that the relation between metabolite and brain development remains stable over time, but the relation between molecules and microbial selection changes at different stages of life. Addressing these limitations in humans is challenging, even if looking at stool metabolites, because overall exposure throughout the gastrointestinal tract is not necessarily reflected in the final concentration of those molecules in stool. Therefore, metabolites from blood plasma could provide more accurate systemic concentrations of molecules, but challenges remain on how to interpret them in humans (69,70).

Given that the VEP is evolutionarily conserved in mammals and can be accurately measured during development, the hypotheses generated in humans in this study are readily testable mechanistically using derived from electroencephalography *in vivo* models in future research. For example, VEP could be assessed in germ-free or defined-microbiome animals (71,72) and may be supplemented with specific molecules such as SCFAs, or colonized with microbial species lacking or providing specific metabolic pathways. Furthermore, molecule concentrations in tissues from the gut to the brain can be directly assessed in these models. Uncovering relations between microbial metabolism and specific molecules may also generate hypotheses that can be confirmed in human data. This study, therefore, provides a foundation for deep investigation of the link between the human gut microbiome and brain development.

## Data Availability

All data produced in the present study are available upon reasonable request to the authors. Upon publication in a peer reviewed journal, data and code will be archived at Dryad and Zenodo respectively

## Acknowledgments

We would like to extend our thanks to all the caregivers and their infants who generously provided their time and samples for this study. We are also grateful to the dedicated nurses and researchers who recruited participants and collected data, ensuring the success of this project. The Khula South African Data Collection Team is Layla Bradford, Simone Williams, Lauren Davel, Tembeka Mhlakwaphalwa, Bokang Methola, Khanyisa Nkubungu, Candice Knipe, Zamazimba Madi, and Nwabisa Mlandu. This research was supported by the Wellcome LEAP 1kD program.

## CRediT

Conceptualization: KSB, ETM, KAD, LJGD, VKC

Data curation: KSB, ETM, GFB, FP, SM, MRZ, MM, DH

Formal analysis: KSB, ETM

Funding acquisition: KSB, NP, DCA, DKJ, SCRW, DA, MG, WPF, KAD, LJGD, VKC

Investigation: KSB, ETM, GFB, SM, MRZ, MM, DH

Methodology: KSB, NP, DCA, DKJ, SCRW, DA, MG, WPF, KAD, LJGD, VKC

Project administration: MRZ, MM, DH, LJGD, VKC

Software: KSB, ETM, GFB, CH, LJGD

Resources: SM, AS, FP, MRZ, MM, LD, CB, Khula, KAD, VKC

Supervision: KSB, LJGD, VKC

Validation: KSB, ETM

Visualization: KSB, ETM

Writing – original draft: KSB, ETM, LJGD, VKC

Writing – review & editing: KSB, ETM, CH, NP, KAD

## Supplemental Figures

**Figure S2:**
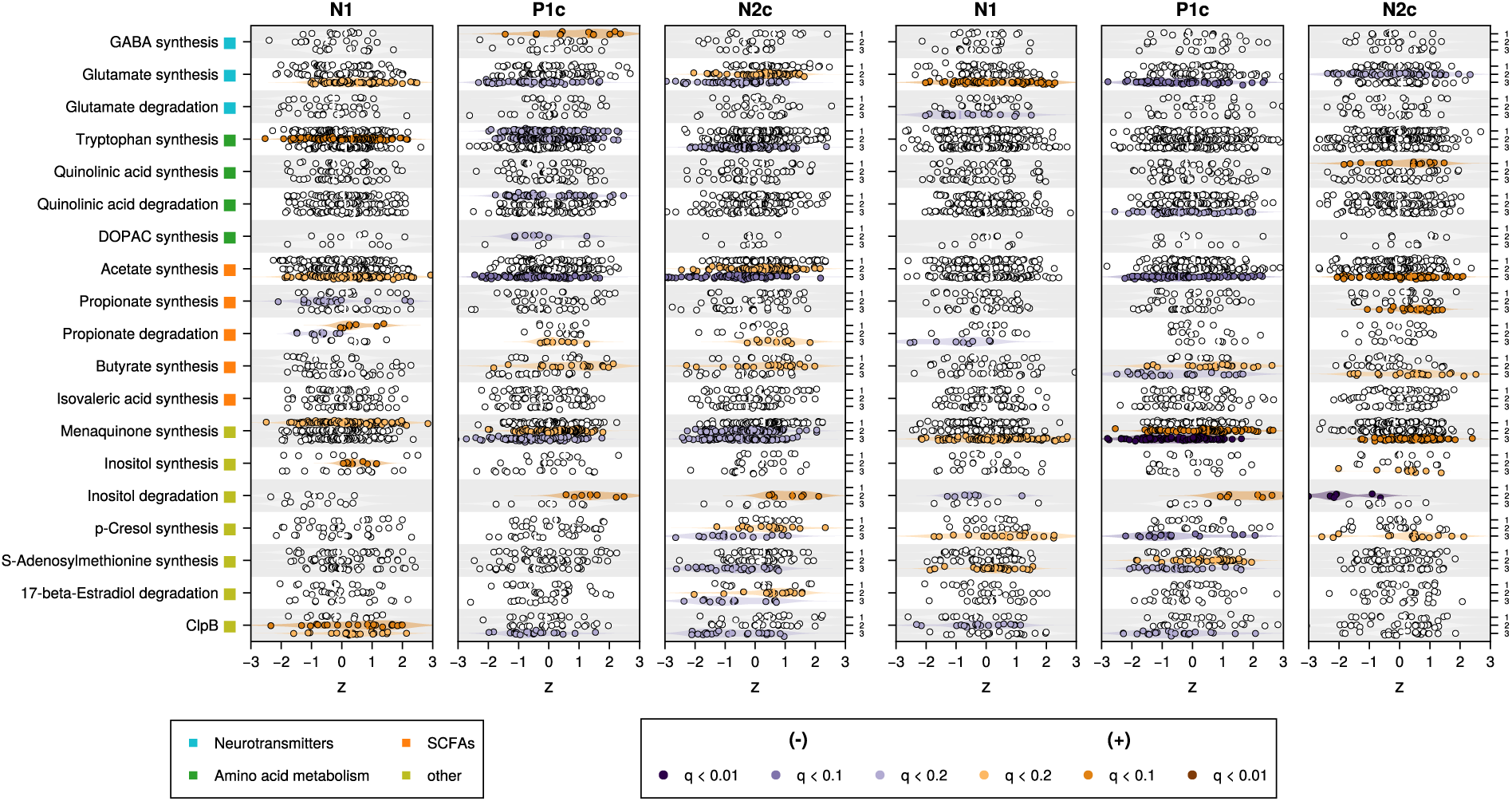
Concurrent feature set enrichment analysis of microbial neuroactive genes and VEP for three visits. FSEA results for all gene sets where at least one visit had a significant hit (q < 0.2) with at least one VEP latency (A) or amplitude (B). Dots indicate the Z-statistic from logistic regression for each gene in a gene set. Vertical bars indicate the median Z-statistic for the gene set as a whole. Y-axis position for each gene set indicates visit number. Visit 1 for inositol degradation and DOPAC synthesis were not tested, since there were fewer than 5 genes from those genesets present in the sample (See Methods).

**Figure S3:**
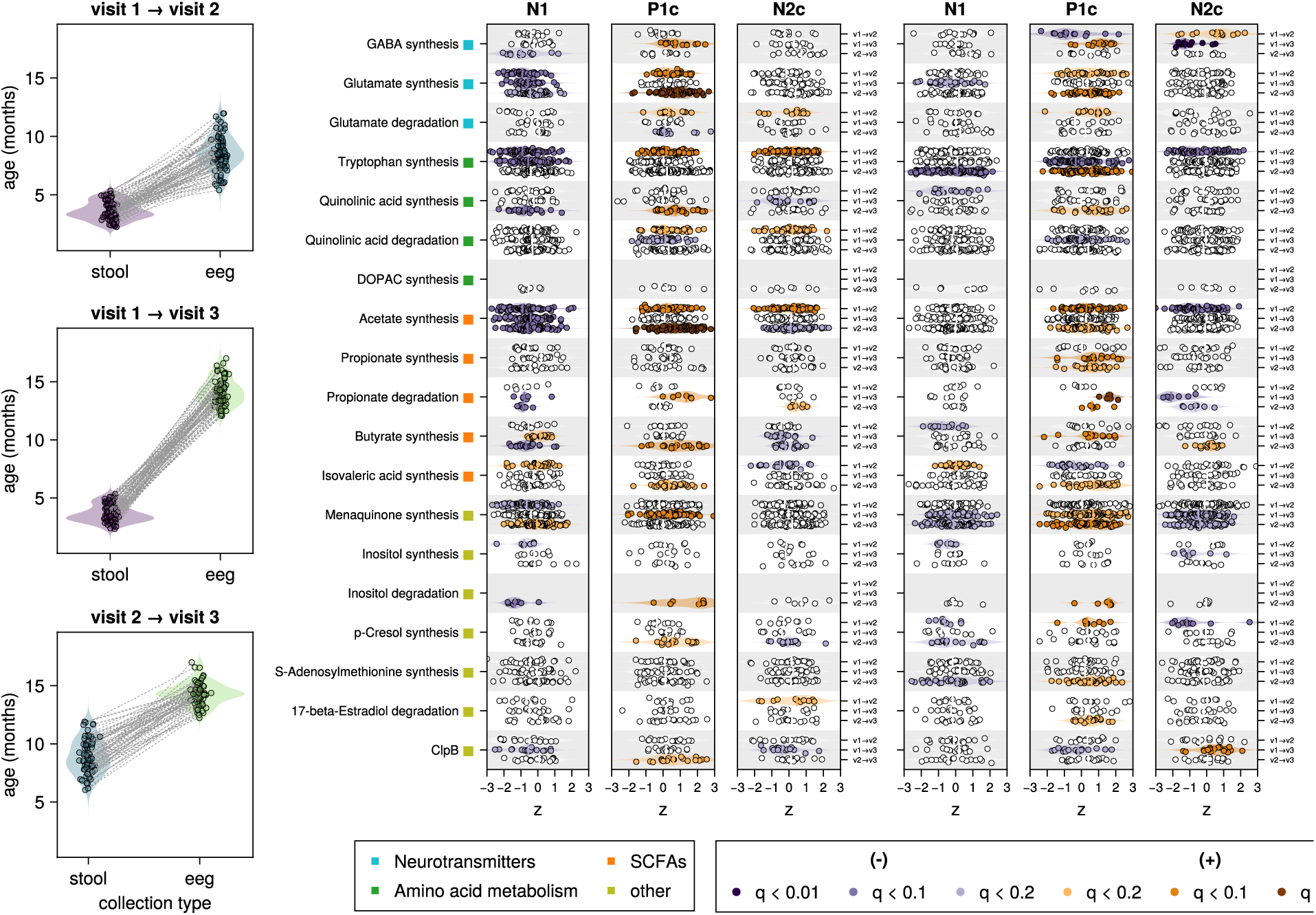
Gut microbial genes predict future VEP latencies and amplitudes. (A) Age distributions for stool samples (left) and VEP (right) for each longitudinal comparison (same individual) tested, V1 stool →V2 VEP, V1 stool →V3 VEP, and V2 stool →V3 VEP. As in Figure S2, (B) and (C) show FSEA results for all genesets where at least one visit had a significant hit (q < 0.2) with at least one VEP latency or amplitude respectively. Dots indicate the Z-statistic from logistic regression for each gene in a gene set. Vertical bars indicate the median Z-statistic for the gene set as a whole. The Y-axis position for each gene set indicates longitudinal comparison. V1 → V2 and V1 → 3 for inositol degradation and DOPAC synthesis were not tested, since there were fewer than 5 genes from those genesets present in the sample (See Methods).

## Bibliography

1. Rhee SH, Pothoulakis C, Mayer EA (2009): Principles and Clinical Implications of the Brain-Gut-Enteric Microbiota Axis. Nature Reviews Gastroenterology & Hepatology 6: 306–314.

2. Collins SM, Surette M, Bercik P (2012): The Interplay between the Intestinal Microbiota and the Brain. Nature Reviews Microbiology 10: 735–742.

3. Brett BE, Weerth C de (2019): The Microbiota–Gut–Brain Axis: A Promising Avenue to Foster Healthy Developmental Outcomes. Developmental Psychobiology 61: 772–782.

4. Gilbert JA, Blaser MJ, Caporaso JG, Jansson JK, Lynch SV, Knight R (2018): Current Understanding of the Human Microbiome. Nature Medicine 24: 392–400.

5. Valles-Colomer M, Falony G, Darzi Y, Tigchelaar EF, Wang J, Tito RY, et al. (2019): The Neuroactive Potential of the Human Gut Microbiota in Quality of Life and Depression. Nature microbiology 4: 623– 632.

6. Janik R, Thomason LAM, Stanisz AM, Forsythe P, Bienenstock J, Stanisz GJ (2016): Magnetic Resonance Spectroscopy Reveals Oral Lactobacillus Promotion of Increases in Brain GABA, N-acetyl Aspartate and Glutamate. NeuroImage 125: 988–995.

7. Ahmed H, Leyrolle Q, Koistinen V, Kärkkäinen O, Layé S, Delzenne N, Hanhineva K (2022): Microbiota-Derived Metabolites as Drivers of Gut–Brain Communication. Gut Microbes 14: 2102878.

8. Matsumoto M, Kibe R, Ooga T, Aiba Y, Sawaki E, Koga Y, Benno Y (2013): Cerebral Low-Molecular Metabolites Influenced by Intestinal Microbiota: A Pilot Study. Frontiers in Systems Neuroscience 7: 9.

9. Kawase T, Nagasawa M, Ikeda H, Yasuo S, Koga Y, Furuse M (2017): Gut Microbiota of Mice Putatively Modifies Amino Acid Metabolism in the Host Brain. The British Journal of Nutrition 117: 775–783.

10. Filpa V, Moro E, Protasoni M, Crema F, Frigo G, Giaroni C (2016): Role of Glutamatergic Neurotransmission in the Enteric Nervous System and Brain-Gut Axis in Health and Disease. Neuropharmacology 111: 14–33.

11. Baj A, Moro E, Bistoletti M, Orlandi V, Crema F, Giaroni C (2019): Glutamatergic Signaling Along The Microbiota-Gut-Brain Axis. International Journal of Molecular Sciences 20: 1482.

12. Agus A, Planchais J, Sokol H (2018): Gut Microbiota Regulation of Tryptophan Metabolism in Health and Disease. Cell Host & Microbe 23: 716–724.

13. Dalile B, Van Oudenhove L, Vervliet B, Verbeke K (2019): The Role of Short-Chain Fatty Acids in Microbiota–Gut–Brain Communication. Nature Reviews Gastroenterology & Hepatology 16: 461– 478.

14. Erny D, Dokalis N, Mezö C, Castoldi A, Mossad O, Staszewski O, et al. (2021): Microbiota-Derived Acetate Enables the Metabolic Fitness of the Brain Innate Immune System during Health and Disease. Cell Metabolism 33: 2260–2276.

15. Parker A, Fonseca S, Carding SR (2019): Gut Microbes and Metabolites as Modulators of Blood-Brain Barrier Integrity and Brain Health. Gut Microbes 11: 135–157.

16. Aburto MR, Cryan JF (2024): Gastrointestinal and Brain Barriers: Unlocking Gates of Communication across the Microbiota–Gut–Brain Axis. Nature Reviews Gastroenterology & Hepatology 21: 222–247.

17. Bonham K, Bottino GF, McCann SH, Beauchemin J, Weisse E, Barry F, et al. (2023): Gut-Resident Microorganisms and Their Genes Are Associated with Cognition and Neuroanatomy in Children. Science Advances 9: eadi497.

18. Lippé S, Roy M-S, Perchet C, Lassonde M (2007): Electrophysiological Markers of Visuocortical Development. Cerebral Cortex 17: 100–107.

19. Dean DC, O’Muircheartaigh J, Dirks H, Waskiewicz N, Lehman K, Walker L, et al. (2014): Modeling Healthy Male White Matter and Myelin Development: 3 through 60 ∼ Months of Age. NeuroImage 84: 742–752.

20. Ahrens AP, Hyötyläinen T, Petrone JR, Igelström K, George CD, Garrett TJ, et al. (2024): Infant Microbes and Metabolites Point to Childhood Neurodevelopmental Disorders. Cell 187: 1853–1873.

21. Meyer K, Lulla A, Debroy K, Shikany JM, Yaffe K, Meirelles O, Launer LJ (2022): Association of the Gut Microbiota With Cognitive Function in Midlife. JAMA Network Open 5: e2143941.

22. Callaghan BL, Fields A, Gee DG, Gabard-Durnam L, Caldera C, Humphreys KL, et al. (2020): Mind and Gut: Associations between Mood and Gastrointestinal Distress in Children Exposed to Adversity. Development and Psychopathology 32: 309–328.

23. Canipe LG, Sioda M, Cheatham CL (2021): Diversity of the Gut-Microbiome Related to Cognitive Behavioral Outcomes in Healthy Older Adults. Archives of Gerontology and Geriatrics 96: 104464.

24. Lupori L, Cornuti S, Mazziotti R, Borghi E, Ottaviano E, Cas MD, et al. (2022): The Gut Microbiota of Environmentally Enriched Mice Regulates Visual Cortical Plasticity. Cell Reports 38. 10.1016/j.celrep.2021.110212

25. Deen B, Richardson H, Dilks DD, Takahashi A, Keil B, Wald LL, et al. (2017): Organization of High-Level Visual Cortex in Human Infants. Nature Communications 8: 13995.

26. Ellis CT, Yates TS, Skalaban LJ, Bejjanki VR, Arcaro MJ, Turk-Browne NB (2021): Retinotopic Organization of Visual Cortex in Human Infants. Neuron 109: 2616–2626.

27. Kiorpes L (2015): Visual Development in Primates: Neural Mechanisms and Critical Periods. Developmental Neurobiology 75: 1080–1090.

28. Gabard-Durnam L, McLaughlin KA (2020): Sensitive Periods in Human Development: Charting a Course for the Future. Current Opinion in Behavioral Sciences 36: 120–128.

29. Margolis ET, Davel L, Bourke NJ, Bosco C, Zieff MR, Monachino AD, et al. (2024): Longitudinal Effects of Prenatal Alcohol Exposure on Visual Neurodevelopment over Infancy. Developmental Psychology 60: 1673–1698.

30. Zieff MR, Miles M, Mbale E, Eastman E, Ginnell L, Williams SCR, et al. (2024): Characterizing Developing Executive Functions in the First 1000 Days in South Africa and Malawi: The Khula Study. Wellcome Open Research 9: 157.

31. Mlandu N, McCormick SA, Davel L, Zieff MR, Bradford L, Herr D, et al. (2024): Evaluating a Novel High-Density EEG Sensor Net Structure for Improving Inclusivity in Infants with Curly or Tightly Coiled Hair. Developmental Cognitive Neuroscience 67: 101396.

32. Monachino AD, Lopez KL, Pierce LJ, Gabard-Durnam LJ (2022): The HAPPE plus Event-Related (HAPPE+ER) Software: A Standardized Preprocessing Pipeline for Event-Related Potential Analyses. Modeling Neural Development 57: 101140.

33. Comeau AM, Filloramo GV (2023): Preparing Multiplexed WGS/MetaG Libraries with the Illumina DNA Prep Kit for the Illumina NextSeq or MiSeq. Retrieved August 14, 2024, from https://www.protocols.io/view/preparing-multiplexed-wgs-metag-libraries-with-the-b5z4q78w

34. Bates D, Mächler M, Bolker B, Walker S (2015): Fitting Linear Mixed-Effects Models Using Lme4. Journal of Statistical Software 67: 1–48.

35. Beghini F, McIver LJ, Blanco-Míguez A, Dubois L, Asnicar F, Maharjan S, et al. (2021): Integrating Taxonomic, Functional, and Strain-Level Profiling of Diverse Microbial Communities with bioBakery 3. Elife 10. 10.7554/eLife.65088

36. Bezanson J, Edelman A, Karpinski S, Shah VB (2017): Julia: A Fresh Approach to Numerical Computing. Siam Review 59: 65–98.

37. Bonham K, Kayisire A, Luo A, Klepac-Ceraj V (2021): Microbiome.Jl and BiobakeryUtils.Jl - Julia Packages for Working with Microbial Community Data. Journal of Open Source Software 6: 3876.

38. Suzek BE, Huang H, McGarvey P, Mazumder R, Wu CH (2007): UniRef: Comprehensive and Non-Redundant UniProt Reference Clusters. Bioinformatics 23: 1282–1288.

39. Kanehisa M, Goto S, Kawashima S, Okuno Y, Hattori M (2004): The KEGG Resource for Deciphering the Genome. Nucleic Acids Research 32: D277–D280.

40. Subramanian A, Tamayo P, Mootha VK, Mukherjee S, Ebert BL, Gillette MA, et al. (2005): Gene Set Enrichment Analysis: A Knowledge-Based Approach for Interpreting Genome-Wide Expression Profiles. Proceedings of the National Academy of Sciences 102: 15545–15550.

41. Bonham K (2024, September 1): Klepac-Ceraj-Lab/EEGMicrobiome: Initial Submission. 10.5281/zenodo.13625448

42. Yassour M, Vatanen T, Siljander H, Hämäläinen A-M, Härkönen T, Ryhänen SJ, et al. (2016): Natural History of the Infant Gut Microbiome and Impact of Antibiotic Treatment on Bacterial Strain Diversity and Stability. Science Translational Medicine 8: 343.

43. Koenig JE, Spor A, Scalfone N, Fricker AD, Stombaugh J, Knight R, et al. (2011): Succession of Microbial Consortia in the Developing Infant Gut Microbiome. Proceedings of the National Academy of Sciences of the United States of America 4578–4585.

44. Bäckhed F, Roswall J, Peng Y, Feng Q, Jia H, Kovatcheva-Datchary P, et al. (2015): Dynamics and Stabilization of the Human Gut Microbiome during the First Year of Life. Cell host & microbe 17: 690– 703.

45. Li J, Lin JC, Wang H, Peterson JW, Furie BC, Furie B, et al. (2003): Novel Role of Vitamin K in Preventing Oxidative Injury to Developing Oligodendrocytes and Neurons. Journal of Neuroscience 23: 5816–5826.

46. Lippe S, Kovacevic N, McIntosh R (2009): Differential Maturation of Brain Signal Complexity in the Human Auditory and Visual System. Frontiers in Human Neuroscience 3. 10.3389/neuro.09.048.2009

47. Carlson AL, Xia K, Azcarate-Peril MA, Goldman BD, Ahn M, Styner MA, et al. (2018): Infant Gut Microbiome Associated With Cognitive∼Development. Biological Psychiatry 83: 148–159.

48. Fagiolini M, Hensch TK (2000): Inhibitory Threshold for Critical-Period Activation in Primary Visual Cortex. Nature 404: 183–186.

49. Hensch TK, Fagiolini M, Mataga N, Stryker MP, Baekkeskov S, Kash SF (1998): Local GABA Circuit Control of Experience-Dependent Plasticity in Developing Visual Cortex. Science 282: 1504–1508.

50. Takesian AE, Bogart LJ, Lichtman JW, Hensch TK (2018): Inhibitory Circuit Gating of Auditory Critical-Period Plasticity. Nature Neuroscience 21: 218–227.

51. Takanaga H, Ohtsuki S, Hosoya Ki n, Terasaki T (2001): GAT2/BGT-1 as a System Responsible for the Transport of Gamma-Aminobutyric Acid at the Mouse Blood-Brain Barrier. Journal of Cerebral Blood Flow and Metabolism: Official Journal of the International Society of Cerebral Blood Flow and Metabolism 21: 1232–1239.

52. Al-Sarraf H (2002): Transport of 14C-γ-aminobutyric Acid into Brain, Cerebrospinal Fluid and Choroid Plexus in Neonatal and Adult Rats. Developmental Brain Research 139: 121–129.

53. Shyamaladevi N, Jayakumar AR, Sujatha R, Paul V, Subramanian EH (2002): Evidence That Nitric Oxide Production Increases γ-Amino Butyric Acid Permeability of Blood-Brain Barrier. Brain Research Bulletin 57: 231–236.

54. Vetencourt JFM, Tiraboschi E, Spolidoro M, Castrén E, Maffei L (2011): Serotonin Triggers a Transient Epigenetic Mechanism That Reinstates Adult Visual Cortex Plasticity in Rats. European Journal of Neuroscience 33: 49–57.

55. Gu Q, Singer W (1995): Involvement of Serotonin in Developmental Plasticity of Kitten Visual Cortex. European Journal of Neuroscience 7: 1146–1153.

56. Lugo-Huitrón R, Ugalde Muñiz P, Pineda B, Pedraza-Chaverrí J, Ríos C, Cruz V Pérez-de la (2013): Quinolinic Acid: An Endogenous Neurotoxin with Multiple Targets. Oxidative Medicine and Cellular Longevity 2013: 104024.

57. Tavares RG, Tasca CI, Santos CES, Wajner M, Souza DO, Dutra-Filho CS (2000): Quinolinic Acid Inhibits Glutamate Uptake into Synaptic Vesicles from Rat Brain. NeuroReport 11: 249.

58. Hong SZ, Mesik L, Grossman CD, Cohen JY, Lee B, Severin D, et al. (2022): Norepinephrine Potentiates and Serotonin Depresses Visual Cortical Responses by Transforming Eligibility Traces. Nature Communications 13: 3202.

59. Yano JM, Yu K, Donaldson GP, Shastri GG, Ann P, Ma L, et al. (2015): Indigenous Bacteria from the Gut Microbiota Regulate Host Serotonin Biosynthesis. Cell 161: 264–276.

60. Jameson KG, Hsiao EY (2018): Linking the Gut Microbiota to a Brain Neurotransmitter. Trends in Neurosciences 41: 413–414.

61. Takesian AE, Hensch TK (2013): Balancing Plasticity/Stability across Brain Development. Progress in Brain Research 207: 3–34.

62. Madhavarao CN, Arun P, Moffett JR, Szucs S, Surendran S, Matalon R, et al. (2005): Defective N-acetylaspartate Catabolism Reduces Brain Acetate Levels and Myelin Lipid Synthesis in Canavan’s Disease. Proceedings of the National Academy of Sciences 102: 5221–5226.

63. Chen T, Noto D, Hoshino Y, Mizuno M, Miyake S (2019): Butyrate Suppresses Demyelination and Enhances Remyelination. Journal of Neuroinflammation 16: 165.

64. Grüter T, Mohamad N, Rilke N, Blusch A, Sgodzai M, Demir S, et al. (2023): Propionate Exerts Neuroprotective and Neuroregenerative Effects in the Peripheral Nervous System. Proceedings of the National Academy of Sciences of the United States of America 120. 10.1073/pnas.2216941120

65. Caetano-Silva ME, Rund L, Hutchinson NT, Woods JA, Steelman AJ, Johnson RW (2023): Inhibition of Inflammatory Microglia by Dietary Fiber and Short-Chain Fatty Acids. Scientific Reports 13: 2819.

66. Barnikol UB, Amunts K, Dammers J, Mohlberg H, Fieseler T, Malikovic A, et al. (2006): Pattern Reversal Visual Evoked Responses of V1/V2 and V5/MT as Revealed by MEG Combined with Probabilistic Cytoarchitectonic Maps. NeuroImage 31: 86–108.

67. You Y, Klistorner A, Thie J, Graham SL (2011): Latency Delay of Visual Evoked Potential Is a Real Measurement of Demyelination in a Rat Model of Optic Neuritis. Investigative Ophthalmology & Visual Science 52: 6911–6918.

68. Gao W, Salzwedel AP, Carlson AL, Xia K, Azcarate-Peril MA, Styner MA, et al. (2019): Gut Microbiome and Brain Functional Connectivity in Infants-a Preliminary Study Focusing on the Amygdala. Psychopharmacology 236: 1641–1651.

69. Deng K, Xu J-j, Shen L, Zhao H, Gou W, Xu F, et al. (2023): Comparison of Fecal and Blood Metabolome Reveals Inconsistent Associations of the Gut Microbiota with Cardiometabolic Diseases. Nature Communications 14: 571.

70. Dekkers KF, Sayols-Baixeras S, Baldanzi G, Nowak C, Hammar U, Nguyen D, et al. (2022): An Online Atlas of Human Plasma Metabolite Signatures of Gut Microbiome Composition. Nature Communications 13: 5370.

71. Wymore Brand M, Wannemuehler MJ, Phillips GJ, Proctor A, Overstreet A-M, Jergens AE, et al. (2015): The Altered Schaedler Flora: Continued Applications of a Defined Murine Microbial Community. ILAR Journal 56: 169–178.

72. Kennedy EA, King KY, Baldridge MT (2018): Mouse Microbiota Models: Comparing Germ-Free Mice and Antibiotics Treatment as Tools for Modifying Gut Bacteria. Frontiers in Physiology 9. 10.3389/fphys.2018.01534

