## Supplementary material for "Co-development of gut microbial metabolism and visual neural circuitry over human infancy": Table S6

| EEG Quality Control Variables | | | |
| --- | --- | --- | --- |
|  | **Visit 1 EEG (N=97)** | **Visit 2 EEG (N=130)** | **Visit 3 EEG (N=131)** |
| **VEP Trial Retention** |  |  |  |
| **Number of Collected VEP Trials** |  |  |  |
| Mean (SD) | 100 (0) | 100 (0) | 100 (0) |
| Median [Min, Max] | 100 [100, 100] | 100 [100, 100] | 100 [100, 100] |
| **Number of Retained VEP Trials** |  |  |  |
| Mean (SD) | 96.0 (9.12) | 96.5 (11.3) | 98.0 (6.35) |
| Median [Min, Max] | 100 [56.0, 100] | 100 [21.0, 100] | 100 [58.0, 100] |
| **Region of Interest (ROI) Channel Retention** | |  |  |
| **Number of Retained Channels in ROI** | |  |  |
| Mean (SD) | 4.45 (0.817) | 4.18 (1.03) | 4.20 (0.898) |
| Median [Min, Max] | 5.00 [1.00, 5.00] | 5.00 [1.00, 5.00] | 4.00 [1.00, 5.00] |
| **Correlation of Data Pre-v s. Post- Wavelet Thresholding (Pearson’s r)** | | |  |
| **At 5 Hz** |  |  |  |
| Mean (SD) | 0.499 (0.171) | 0.403 (0.242) | 0.471 (0.275) |
| Median [Min, Max] | 0.531 [0.0389, 0.875] | 0.411 [0.0180, 0.869] | 0.521 [0.00532, 0.947] |
| **At 8 Hz** |  |  |  |
| Mean (SD) | 0.409 (0.160) | 0.381 (0.234) | 0.494 (0.283) |
| Median [Min, Max] | 0.416 [0.0689, 0.776] | 0.383 [0.0240, 0.849] | 0.562 [0.0146, 0.931] |
| **At 12 Hz** |  |  |  |
| Mean (SD) | 0.384 (0.158) | 0.320 (0.219) | 0.396 (0.258) |
| Median [Min, Max] | 0.394 [0.0377, 0.757] | 0.310 [0.0159, 0.765] | 0.447 [0.0196, 0.857] |
| **At 20 Hz** |  |  |  |
| Mean (SD) | 0.457 (0.154) | 0.379 (0.237) | 0.436 (0.270) |
| Median [Min, Max] | 0.467 [0.0839, 0.738] | 0.410 [0.0157, 0.830] | 0.524 [0.0162, 0.841] |
| Note. EEG data were pre-processed and VEPs were extracted using HAPPE+ER v3.3 software, an automated open-source EEG processing software validated for infant data (Monachino et al., 2022). | | | |
