## Supplementary material for "Co-development of gut microbial metabolism and visual neural circuitry over human infancy": Table S5

**Table X**

*HAPPE v3.3 GenerateERPs Script Parameters*

| **Average or Individual Trials** | Average |
| --- | --- |
| **Channels of Interest** | E70, E71, E75, E76, E83 |
| **Bad Channels Included/Excluded** | Included |
| **Calculating ERP Values** | On |
| **Windows** | Visit 1 & 2:  Min 40-100 milliseconds  Max 75-175 milliseconds  Min 100-325 milliseconds  Visit 3:  Min 35-80 milliseconds  Max 75-130 milliseconds  Min 100-275 milliseconds |
