## Supplementary material for "Co-development of gut microbial metabolism and visual neural circuitry over human infancy": Table S4

**Table X**

*HAPPE v3.3 Pre-Processing Script Parameters*

| **Density** | High (>30 channels) |
| --- | --- |
| **Resting State or Task** | Task |
| **ERP Analysis** | Yes |
| **Acquisition Layout** | 128 channel EGI HydroCel Geodesic Sensor Net |
| **Channels** | All except E1, E8, E14, E17, E21, E25, E32, E38, E43, E44, E48, E49, E56, E63, E68, E73, E81, E88, E94, E99, E107, E113, E114, E119, E120, E121, E125, E126, E127, E128 |
| **Line Noise** |  |
| **Line Noise Frequency** | 50 Hz |
| **Line Noise Reduction Method** | CleanLine - Default |
| **Resample** | Off |
| **Filter** |  |
| **Filter - Lowpass Cutoff** | 30 Hz |
| **Filter - Highpass Cutoff** | 0.3 Hz |
| **Filter Type** | EEGLAB’s FIR |
| **Bad Channel Detection** | On |
| **Bad Channel Detection Method** | Default |
| **Wavelet Thresholding** | Default |
| **Wavelet Threshold Rule** | Hard |
| **MuscIL** | Off |
| **Segmentation** | On |
| **Starting Parameter for Stimulus** | - 0.1 seconds |
| **Ending Parameter for Stimulus** | 0.5 seconds |
| **Task Offset** | 11 milliseconds |
| **Baseline Correction** | On |
| **Baseline Correction Start** | - 100 milliseconds |
| **Baseline Correction End** | 0 milliseconds |
| **Interpolation** | Off |
| **Segment Rejection** | On |
| **Segment Rejection Method** | Amplitude criteria only |
| **Minimum Segment Rejection Threshold** | - 200 |
| **Maximum Segment Rejection Threshold** | 200 |
| **Segment Rejection based on All Channels or ROI** | ROI |
| **ROI Channels** | E70, E71, E75, E76, E83 |
| **Re-Reference Method** | Average |
