## Supplementary material for "Co-development of gut microbial metabolism and visual neural circuitry over human infancy": Table S3

| Predictive Analyses Demographic Information | | | |
| --- | --- | --- | --- |
|  | **Visit 1 🡪 Visit 2 Analysis Cohort (N=84)** | **Visit 1 🡪 Visit 3 Analysis Cohort (N=76)** | **Visit 2 🡪 Visit 3 Analysis Cohort (N=69)** |
| **Maternal Place of Birth** |  |  |  |
| South Africa | 83 (98.8%) | 76 (100%) | 69 (100%) |
| In the African Continent (not South Africa) | 1 (1.2%) | 0 (0%) | 0 (0%) |
| **Primary Spoken Language** |  |  |  |
| Xhosa Language | 79 (94.0%) | 72 (94.7%) | 66 (95.7%) |
| Sotho Language | 2 (2.4%) | 2 (2.6%) | 1 (1.4%) |
| Zulu Language | 1 (1.2%) | 1 (1.3%) | 0 (0%) |
| English Language | 0 (0%) | 0 (0%) | 1 (1.4%) |
| Ndebele Language | 1 (1.2%) | 0 (0%) | 0 (0%) |
| Afrikaans Language | 1 (1.2%) | 1 (1.3%) | 1 (1.4%) |
| **Maternal Age at Infant Birth (years)** |  |  |  |
| Mean (SD) | 29.7 (5.42) | 29.9 (5.55) | 29.1 (5.66) |
| Median [Min, Max] | 30.0 [18.0, 40.0] | 30.0 [18.0, 40.0] | 29.0 [18.0, 41.0] |
| Missing | 0 (0%) | 0 (0%) | 0 (0%) |
| **Maternal Educational Attainmentᵃ** |  |  |  |
| Completed Grade 6 (Standard 4) to Grade 7 (Standard 5) | 2 (2.4%) | 1 (1.3%) | 2 (2.9%) |
| Completed Grade 8 (Standard 6) to Grade 11 (Standard 9) i.e., high school without matriculating | 33 (39.3%) | 33 (43.4%) | 27 (39.1%) |
| Completed Grade 12 (Standard 10) i.e., high school | 38 (45.2%) | 31 (40.8%) | 30 (43.5%) |
| Part of university/ college/ post-matric education | 6 (7.1%) | 4 (5.3%) | 5 (7.2%) |
| Completed university/ college/ post-matric education | 5 (6.0%) | 7 (9.2%) | 5 (7.2%) |
| **Maternal Monthly Incomeᵇ (South African Rand/ZAR)** |  |  |  |
| Less than R1000 per month | 39 (46.4%) | 39 (51.3%) | 39 (56.5%) |
| R1000 - R5000 per month | 36 (42.9%) | 25 (32.9%) | 20 (29.0%) |
| R5000 - R10,000 per month | 8 (9.5%) | 10 (13.2%) | 6 (8.7%) |
| More than R10,000 per month | 0 (0%) | 0 (0%) | 0 (0%) |
| Unknown | 1 (1.2%) | 2 (2.6%) | 4 (5.8%) |
| **Infant Biological Sex** |  |  |  |
| Female | 38 (45.2%) | 35 (46.1%) | 31 (44.9%) |
| Male | 46 (54.8%) | 41 (53.9%) | 38 (55.1%) |
| ᵃ The South African Educational System was formerly divided into years called standards, similarly to the way the United States Educational System is divided into grades. The equivalent in terms of standards is provided in parentheses next to each mentioned grade. “University/College/Post-Matric Education” refers to tertiary or post-secondary education as defined by the World Bank.  ᵇ At the time of writing (1/16/24), 1 US Dollar = 18.87 South African Rand (ZAR). | | | |
