## Supplementary material for "Co-development of gut microbial metabolism and visual neural circuitry over human infancy": Table S1

**Table S1: Age-Related Changes in VEP Features**

| Model | *b* | *SE* | *p* |
| --- | --- | --- | --- |
| N1 Peak Amplitude Model |  |  |  |
| Age (months) | -0.07 | 0.03 | .036 |
| Number of Retained Trials | 0.03 | 0.02 | .033 |
| P1 Peak Amplitude Model |  |  |  |
| Age (months) | -0.50 | 0.05 | <.001 |
| Number of Retained Trials | -0.03 | 0.03 | .191 |
| N2 Peak Amplitude Model |  |  |  |
| Age (months) | 0.52 | 0.08 | <.001 |
| Number of Retained Trials | 0.01 | 0.04 | .737 |
| N1 Peak Latency Model |  |  |  |
| Age (months) | -0.79 | 0.08 | <.001 |
| Number of Retained Trials | -0.04 | 0.04 | .342 |
| P1 Peak Latency Model |  |  |  |
| Age (months) | -1.21 | 0.12 | <.001 |
| Number of Retained Trials | 0.01 | 0.06 | .842 |
| N2 Peak Latency Model |  |  |  |
| Age (months) | -4.10 | 0.34 | <.001 |
| Number of Retained Trials | 0.06 | 0.16 | .687 |

Six linear mixed models with each VEP feature as the outcome, age in months as the predictor of interest and number of retained trials as a covariate were run using the *lme4* package (Bates et al., 2015) in R to quantify age-related changes in VEP features. P-values were calculated using the *lmerTest* package (Kuznetsova et al., 2017) in R which uses Satterthwaites approximations. The P1 and N2 amplitudes and latencies are corrected for earlier component amplitudes/latencies.
